# An intermediate effect size variant in *UMOD* confers risk for chronic kidney disease

**DOI:** 10.1101/2021.09.27.21263789

**Authors:** Eric Olinger, Céline Schaeffer, Kendrah Kidd, Yurong Cheng, Inès Dufour, Guglielmo Schiano, Holly Mabillard, Elena Pasqualetto, Elhussein A. E. Elhassan, Patrick Hofmann, Daniel G. Fuster, Andreas D. Kistler, Peter J. Conlon, Ian J. Wilson, Stanislav Kmoch, Genomics England Research Consortium, Kai-Uwe Eckardt, Anthony J. Bleyer, Anna Köttgen, Michael Wiesener, John A. Sayer, Luca Rampoldi, Olivier Devuyst

## Abstract

The kidney-specific gene *UMOD* encodes for uromodulin, the most abundant protein excreted in normal urine. Rare, large-effect variants in *UMOD* cause autosomal dominant tubulointerstitial kidney disease (ADTKD) while common, low-effect variants strongly associate with kidney function and risk of chronic kidney disease (CKD) in the general population. It is unknown whether intermediate-effect variants in *UMOD* contribute to CKD. Here, candidate intermediate-effect *UMOD* variants were identified using large population and ADTKD cohorts. Biological and phenotypical effects were investigated using cell models, *in silico* simulations and international databases and biobanks. Eight *UMOD* missense variants reported in ADTKD are present in gnomAD with MAF ranging from 10^−5^ to 10^−3^. Among them, the missense variant p.Thr62Pro is detected in ∼1/1,000 individuals of European ancestry, shows incomplete penetrance but a high genetic load in familial clusters of CKD and is associated with kidney failure in the 100,000 Genomes Project (OR 3.99; 1.84-8.98) and the UK Biobank (OR 4.12; 1.32-12.85). Compared to canonical ADTKD mutations, the p.Thr62Pro carriers displayed reduced disease severity, with slower progression of CKD, intermediate reduction of urinary UMOD levels, in line with an intermediate trafficking defect *in vitro*. Identification of an intermediate-effect *UMOD* variant completes the spectrum of *UMOD*-associated kidney diseases and provides novel insights into the mechanisms of ADTKD and the genetic architecture of CKD.

**Significance Statement:** The genetic architecture of chronic kidney disease (CKD) remains incompletely understood. Variants in the kidney-specific gene *UMOD* cause autosomal dominant tubulointerstitial kidney disease (ADTKD) and are associated with kidney function and risk of CKD in the general population. Here, we identified an intermediate-effect variant, p.Thr62Pro, detected in ∼1:1,000 individuals of European ancestry, that showed a high genetic load in familial clusters of CKD and was associated with an OR of ∼4 for kidney failure in the 100,000 Genomes Project and the UK Biobank. Compared to canonical ADTKD mutations, p.Thr62Pro carriers displayed reduced disease severity and an intermediate trafficking defect. These findings complete the spectrum of *UMOD*-associated kidney diseases and provide a paradigm for the genetic contribution to CKD.

## Introduction

Chronic kidney disease (CKD) has an estimated global prevalence above 10%, with high burden both at individual and societal levels and limited therapeutic options (1). There is a strong genetic predisposition to CKD, as the heritability of the CKD-defining trait estimated glomerular filtration rate (eGFR) is between 30% and 50% (2, 3). The largest and most recent genome-wide association study (GWAS) for eGFR identified >250 genetic loci and explained nearly 20% of that heritability. Among those, variants in the *UMOD* locus display the largest effect size on eGFR and CKD (4). This effect, thought to be associated with increased *UMOD* expression (5), is consistent across different ethnic groups and is also observed for longitudinal traits including rapid decline of eGFR (6).

The biological relevance of the *UMOD* locus is immediately clear, as it is a kidney-specific gene coding for uromodulin (UMOD), the most abundant protein excreted in the normal urine (2, 7). UMOD, which is produced by the thick ascending limb (TAL) of the loop of Henle, plays multiple roles in blood pressure control and protection against kidney stones and urinary tract infections (7, 8). In TAL cells, UMOD enters the secretory pathway to undergo a complex maturation involving the formation of 24 intramolecular disulphide bonds, a rate-limiting step for its proper apical sorting and release into the tubule lumen (9). Ultrarare (MAF<10^−4^) mutations in *UMOD* cause autosomal dominant tubulointerstitial kidney disease (ADTKD-*UMOD*; MIM #162000), characterized by tubular damage and interstitial fibrosis (10). More than 95% of the *UMOD* mutations associated with ADTKD are missense, often targeting cysteine residues and leading to the formation of UMOD aggregates within the endoplasmic reticulum (ER) (gain-of-toxic function) with a sharp decrease of its excretion in urine. These *UMOD* mutations invariably lead to kidney failure in adulthood with a penetrance of 100% (10, 11). Over 100 mutations in *UMOD* have been associated with ADTKD-*UMOD*, with an overall prevalence of ∼2% in patients with kidney failure, representing one of the most common monogenic kidney diseases (12). *UMOD* is thus implicated at both extremes of the genetic disease spectrum: ultrarare variants with large effect size (ADTKD-*UMOD*) and common (MAF>0.05) variants associated with reduced eGFR and risk of CKD in the general population (13).

The availability of large genomic datasets suggests that the dichotomy of rare, high-effect variants causing Mendelian disorders and frequent, low-effect variants involved in complex diseases must be completed by variants with intermediate effect sizes. These intermediate-effect variants lie on a spectrum based on allelic frequency, biological consequence, and phenotypical readout. They can lead to either non-fully penetrant Mendelian disease or to an oligo/polygenic model modifying disease expressivity (14, 15). The absence of ultrarare and predicted pathogenic variants in the majority of CKD patients (16) and the missing heritability when studying polymorphisms (17) suggests that intermediate-effect variants, not captured by GWAS but amenable to sequencing approaches, may be part of a continuum underlying CKD. Because *UMOD* is involved at both ends of the genetic spectrum, it is a plausible candidate gene for intermediate-effect variants modulating the risk of CKD.

Here, we identified and characterized intermediate-effect variants in *UMOD* contributing to CKD by crossing general population datasets with curated variants reported in ADTKD; analysing biological and phenotypical effect sizes using *in silico* modelling, cell systems, and databases and biobanks; and validating the impact on kidney failure in the 100,000 Genomes Project and UK Biobank (Figure S1).

## Results

### Identification of candidate intermediate-effect variants in UMOD

The *UMOD* gene is implicated at both extremes of a CKD risk continuum (Figure 1A) but *bona fide* high effect variants cannot be more prevalent than the disease they cause. We first assessed the missense variants in *UMOD* reported in the Genome Aggregation Database (gnomAD) (“Controls”) (18) and in the International ADTKD Cohort (11) and the Human Gene Mutation Database (HGMD^®^) (“Cases”). Shared *UMOD* variants should be either benign (falsely associated with disease), or intermediate-effect variants associated with non-fully penetrant or milder, late-onset disease (Figure 1B & Supplementary Information). The majority of *UMOD* variants listed in gnomAD (360/1000, 36%) were missense variants (Figure S2), with an overall allelic frequency (AF) ranging from 2.0×10^−2^ to 3.6×10^−6^ and only 4 variants with an AF exceeding 10^−3^ (Table S1). Most of these missense variants in gnomAD were rare (258 variants with allele counts 3 or less), and 8% among them (30/360) with strong *in silico* pathogenic scores (REVEL scores>0.75) (19) (Table S2). A total of 132 *UMOD* missense variants associated with ADTKD families were included in the International ADTKD Cohort and the HGMD^®^ database (Table S3). As expected, these variants associated with ADTKD were generally ultrarare (MAF<10^−4^ in 129/132, 98%), with most of them (124/132, 94%) being absent from gnomAD. However, eight missense variants (7 in exon 3 and 1 in exon 7) were detected in both populations (Figure 1C & Table 1). These eight variants were identified in 879 individuals in gnomAD, with three being detected relatively frequently (AF 3.5×10^−4^ – 3.3×10^−3^) whereas the five others were considerably rarer (4.1×10^−6^ – 1.4×10^−5^) (Table 1). Since *in silico* predictions and prevalence of these five ultrarare variants remain compatible with high-effect penetrant variants (Table 1 & Table S2), we focused our investigations of potential intermediate-effect variants on the three more frequent *UMOD* missense variants p.Thr62Pro, p.Leu180Val and p.Thr469Met (Table 1). Ages of p.Thr62Pro and p.Thr469Met carriers are consistent with the overall age distribution in gnomAD, suggesting that bias due to recruitment before clinical presentation of ADTKD-*UMOD* is not a concern.

**Figure 1.**
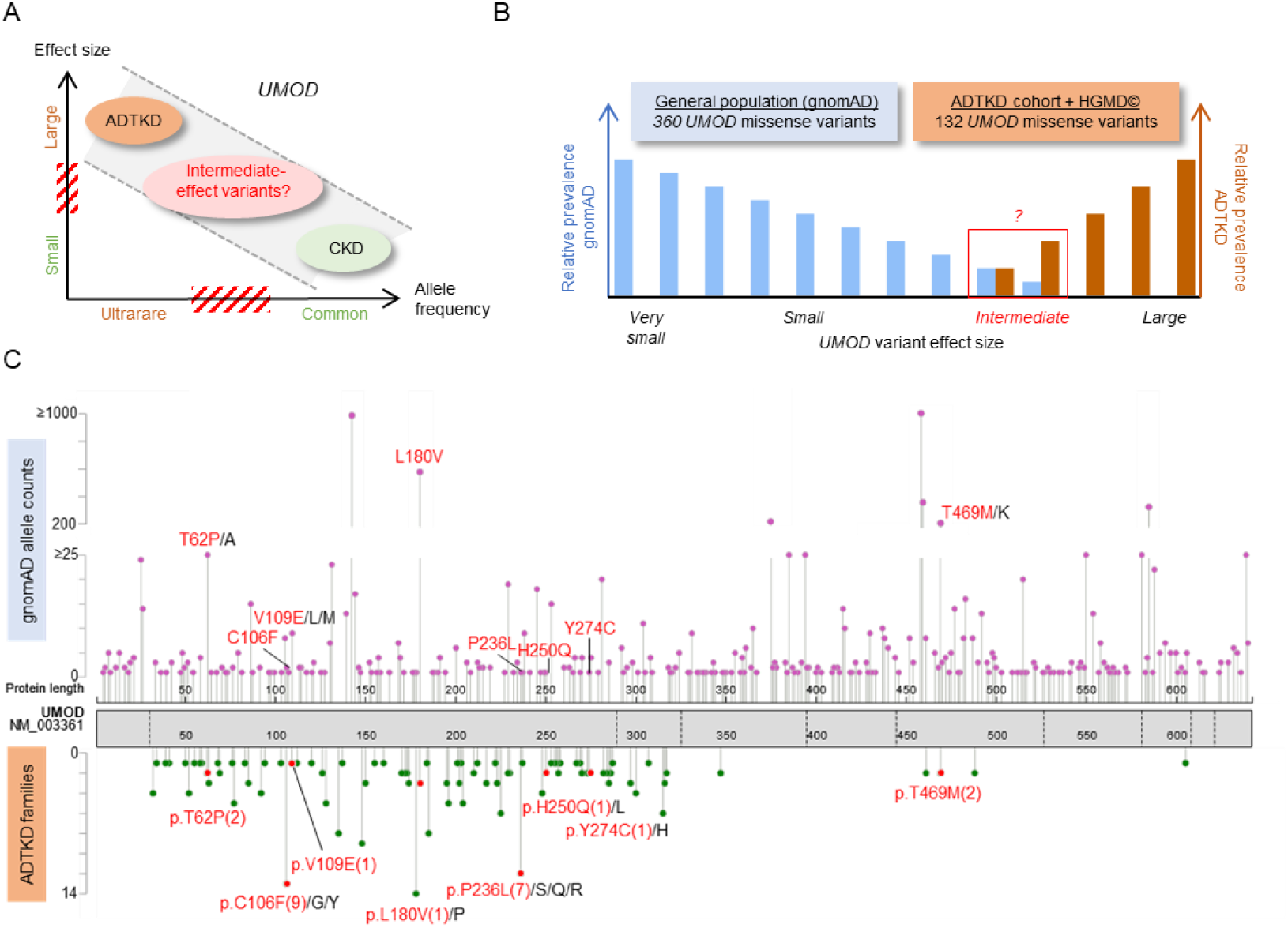
Identification of intermediate-effect *UMOD* variants. (A) Genetic architecture of *UMOD*-associated diseases, showing the continuum of CKD risk associated with ultrarare (AF<10^−4^) high-effect to common (AF>0.05) low-effect *UMOD* variants. Scheme adapted from (17). (B) Schematic representation of study design. 360 distinct missense *UMOD* variants are reported in the genome aggregation database (gnomAD) with an individual allele frequency of 2.0×10^−2^ - 3.6×10^−6^. 132 distinct missense *UMOD* variants reported in ADTKD-*UMOD* have been extracted from the International ADTKD Cohort (726 patients from 585 families) and from The Human Gene Mutation Database (HGMD^®^). The theoretical maximum credible allele frequency of high effect *UMOD* variants (pathogenic) in the overall population, considering the disease prevalence, allelic and genetic heterogeneity and a penetrance of 100% is estimated to be 1×10^−7^ (41) (See Supplementary Information), excluding them (in theory) from gnomAD. Plotted here are the expected relative prevalences of very small to large effect size *UMOD* variants in gnomAD and reported in ADTKD-*UMOD* patients. Conferring non fully penetrant ADTKD or reduced disease expressivity, intermediate-effect *UMOD* variants might be found overlapping at the lower prevalence spectrum in both gnomAD and reported variants in ADTKD-*UMOD*. (C) UMOD protein (NM_003361) in grey with exon boundaries marked by dotted lines. *UMOD* missense variants reported in the gnomAD consortium are plotted on top of the protein with respect to the amino acid position. The y-axis represents gnomAD allele counts. *UMOD* missense variants reported in the international ADTKD cohort and in HGMD^®^ are plotted below the protein. The y-axis represents the number of reported families. Variants that are both reported in gnomAD and in at least 1 ADTKD family are marked in red with family numbers in brackets. Protein and variant visualization using ProteinPaint (42) and cBioPortal MutationMapper (43, 44). Abbreviations: ADTKD, autosomal dominant tubulointerstitial kidney disease; CKD, chronic kidney disease.

**Table 1.**
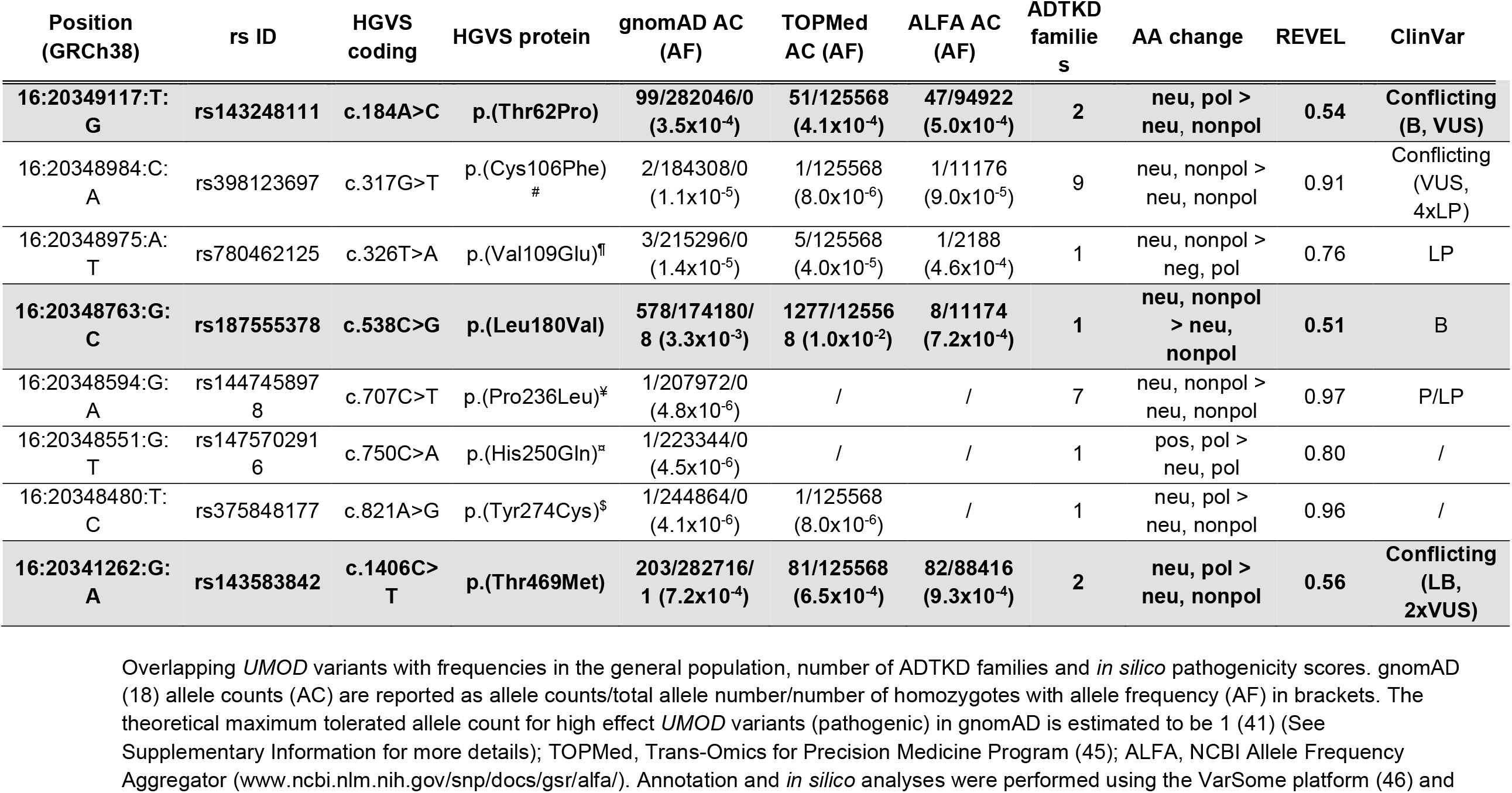
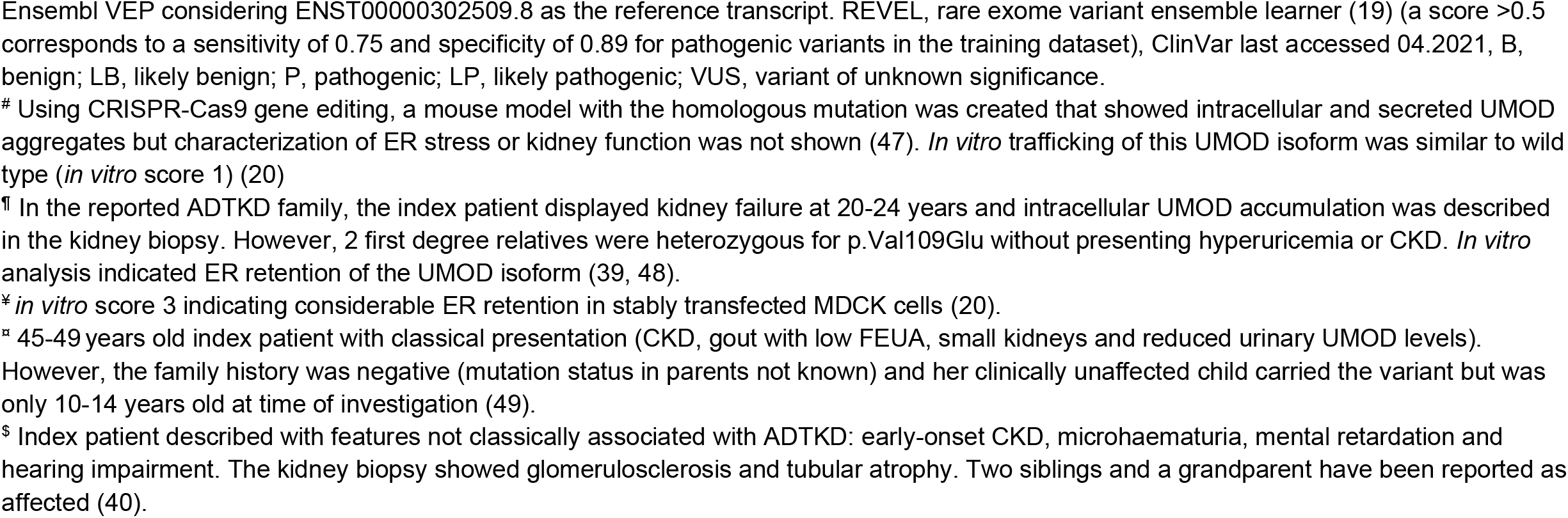
*UMOD* variants detected in gnomAD and ADTKD.

| Position<br>(GRCh38) | rs ID | HGVS<br>coding | HGVS protein | gnomAD AC<br>(AF) | TOPMed<br>AC (AF) | ALFA AC<br>(AF) | ADTKD<br>familie<br>s | AA change | REVEL | ClinVar |
| --- | --- | --- | --- | --- | --- | --- | --- | --- | --- | --- |
| **16:20349117:T:**<br>**G** | **rs143248111** | **c.184A>C** | **p.(Thr62Pro)** | **99/282046/0**<br>**(3.5x10<sup>-4</sup>)** | **51/125568**<br>**(4.1x10<sup>-4</sup>)** | **47/94922**<br>**(5.0x10<sup>-4</sup>)** | **2** | **neu, pol >**<br>**neu, nonpol** | **0.54** | **Conflicting**<br>**(B, VUS)** |
| 16:20348984:C:<br>A | rs398123697 | c.317G>T | p.(Cys106Phe)<br># | 2/184308/0<br>(1.1x10<sup>-5</sup>) | 1/125568<br>(8.0x10<sup>-6</sup>) | 1/11176<br>(9.0x10<sup>-5</sup>) | 9 | neu, nonpol ><br>neu, nonpol | 0.91 | Conflicting<br>(VUS,<br>4xLP) |
| 16:20348975:A:<br>T | rs780462125 | c.326T>A | p.(Val109Glu)<sup>¶</sup> | 3/215296/0<br>(1.4x10<sup>-5</sup>) | 5/125568<br>(4.0x10<sup>-5</sup>) | 1/2188<br>(4.6x10<sup>-4</sup>) | 1 | neu, nonpol ><br>neg, pol | 0.76 | LP |
| **16:20348763:G:**<br>**C** | **rs187555378** | **c.538C>G** | **p.(Leu180Val)** | **578/174180/**<br>**8 (3.3x10<sup>-3</sup>)** | **1277/12556**<br>**8 (1.0x10<sup>-2</sup>)** | **8/11174**<br>**(7.2x10<sup>-4</sup>)** | **1** | **neu, nonpol**<br>**> neu,**<br>**nonpol** | **0.51** | **B** |
| 16:20348594:G:<br>A | rs144745897<br>8 | c.707C>T | p.(Pro236Leu)<sup>¥</sup> | 1/207972/0<br>(4.8x10<sup>-6</sup>) | / | / | 7 | neu, nonpol ><br>neu, nonpol | 0.97 | P/LP |
| 16:20348551:G:<br>T | rs147570291<br>6 | c.750C>A | p.(His250Gln)<sup>²</sup> | 1/223344/0<br>(4.5x10<sup>-6</sup>) | / | / | 1 | pos, pol ><br>neu, pol | 0.80 | / |
| 16:20348480:T:<br>C | rs375848177 | c.821A>G | p.(Tyr274Cys)<sup>\$</sup> | 1/244864/0<br>(4.1x10<sup>-6</sup>) | 1/125568<br>(8.0x10<sup>-6</sup>) | / | 1 | neu, pol ><br>neu, nonpol | 0.96 | / |
| **16:20341262:G:**<br>**A** | **rs143583842** | **c.1406C>**<br>**T** | **p.(Thr469Met)** | **203/282716/**<br>**1 (7.2x10<sup>-4</sup>)** | **81/125568**<br>**(6.5x10<sup>-4</sup>)** | **82/88416**<br>**(9.3x10<sup>-4</sup>)** | **2** | **neu, pol >**<br>**neu, nonpol** | **0.56** | **Conflicting**<br>**(LB,**<br>**2xVUS)** |

Overlapping *UMOD* variants with frequencies in the general population, number of ADTKD families and *in silico* pathogenicity scores. gnomAD (18) allele counts (AC) are reported as allele counts/total allele number/number of homozygotes with allele frequency (AF) in brackets. The theoretical maximum tolerated allele count for high effect *UMOD* variants (pathogenic) in gnomAD is estimated to be 1 (41) (See Supplementary Information for more details); TOPMed, Trans-Omics for Precision Medicine Program (45); ALFA, NCBI Allele Frequency Aggregator (www.ncbi.nlm.nih.gov/snp/docs/gsr/alfa/). Annotation and *in silico* analyses were performed using the VarSome platform (46) and
Ensembl VEP considering ENST00000302509.8 as the reference transcript. REVEL, rare exome variant ensemble learner (19) (a score >0.5 corresponds to a sensitivity of 0.75 and specificity of 0.89 for pathogenic variants in the training dataset), ClinVar last accessed 04.2021, B, benign; LB, likely benign; P, pathogenic; LP, likely pathogenic; VUS, variant of unknown significance.

### p.Thr62Pro UMOD shows intermediate intracellular trafficking defect

The functional consequences of the p.Thr62Pro, p.Thr469Met and p.Leu180Val *UMOD* variants were assessed by monitoring UMOD processing in transiently transfected HEK293 cells. In control conditions, both the immature (100 kDa, “ER-glycosylated”) and mature (120 kDa, “Golgi-glycosylated”) UMOD bands can be detected (Figure 2A). While no or only subtle difference is noted between wild-type, p.Leu180Val and p.Thr469Met UMOD isoforms, p.Thr62Pro UMOD shows an approximately 3-fold increase of the immature/mature ratio compared to wild-type UMOD (Figure 2A). In this setting a typical ADTKD UMOD mutant as p.Cys150Ser is fully retained in the ER, as evidenced by the absence of the mature form, thus demonstrating an intermediate phenotype for p.Thr62Pro isoform between wild-type UMOD and ADTKD mutants. The effect of p.Thr62Pro on protein maturation was confirmed using pulse chase experiments in stably transfected HEK293 cells: p.Thr62Pro UMOD showed an intermediate cellular phenotype between wild-type and p.Cys317Tyr UMOD (a reference ADTKD mutant (20)), with increased amount of immature UMOD at 2h after chase (Figure 2B). Consistent with an intermediate phenotype, colocalization of UMOD with a plasma membrane marker is decreased for p.Thr62Pro compared to wild-type UMOD and to the two variants p.Leu180Val and p.Thr469Met, while it is increased compared with p.Cys150Ser (Figure 2C). Of interest, p.Thr62Pro expression has no evident effect on ER homeostasis at baseline, while the ADTKD mutant p.Cys150Ser induces ER stress, as evidenced by increased expression level of the ER chaperone glucose-regulated protein 78kDa (GRP78, *HSPA5*) and of the spliced isoform of the Unfolded Protein Response (UPR) effector Xbp1 (hXBP1s). However, upon treatment with a low dose of tunicamycin, affecting ER protein folding, cells expressing p.Thr62Pro show increased GRP78 and hXBP1s expression at an intermediate level between wild-type and p.Cys150Ser, suggesting increased ER stress susceptibility (Figure 2D).

**Figure 2.**
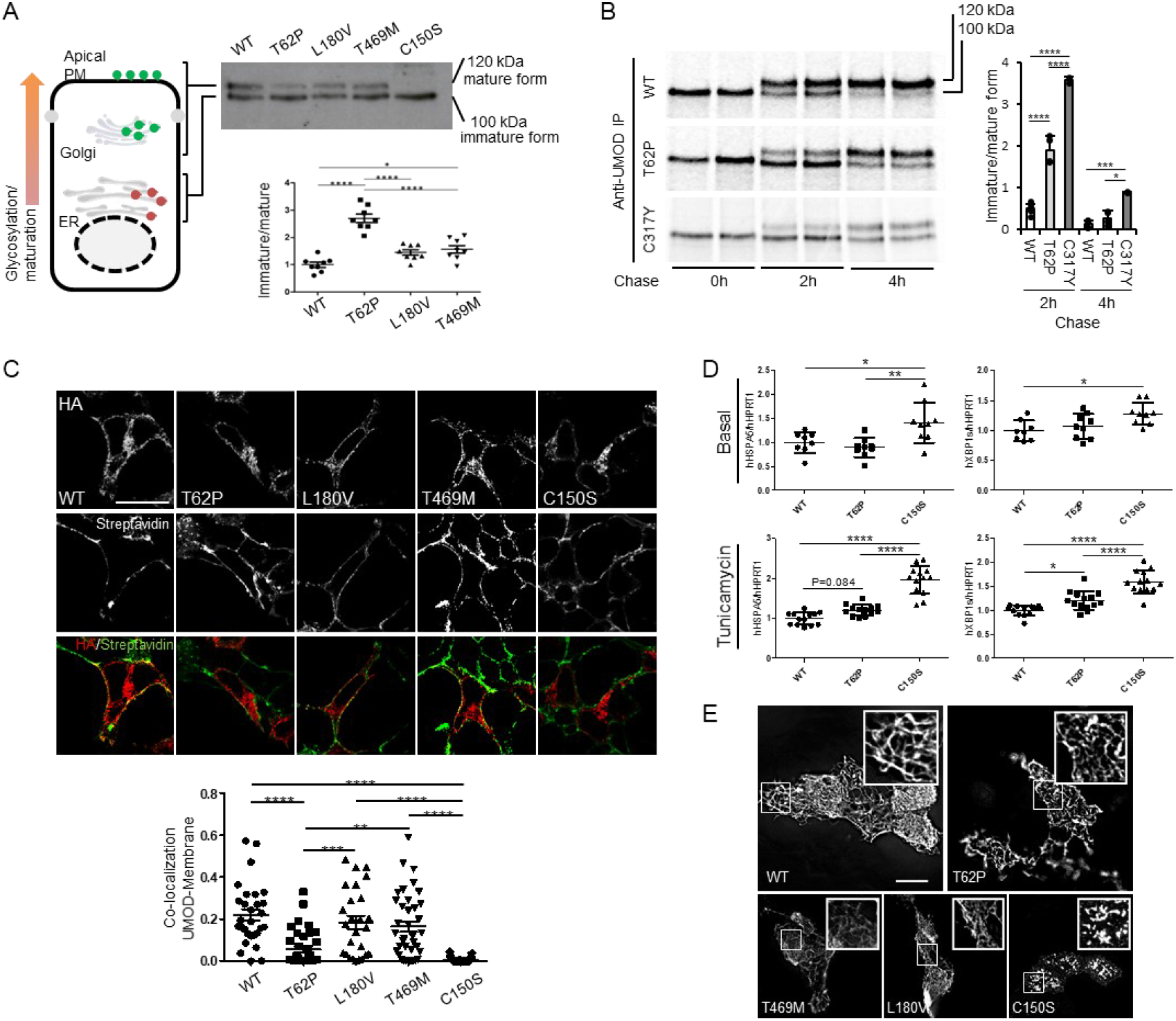
p.Thr62Pro substitution induces intermediate UMOD trafficking and maturation defect. (A) Western blot analysis of UMOD expression in HEK293 cells 6 hours after transfection of the indicated UMOD isoforms. The graph represents the ratio ER glycosylated form (immature) / Golgi glycosylated form (mature). The ratio was not calculated for the p.Cys150Ser mutant as the quantity of mature form is too low to be quantified. N=8 independent experiments; ^*^p<0.05, ^****^p<0.0001 using one way ANOVA, followed by Bonferroni’s multiple comparison test post hoc test. (B) Pulse chase experiments performed in HEK293 cells stably expressing the indicated UMOD isoform. The maturation to fully glycosylated protein is delayed for p.Thr62Pro isoform compared to the wild type one, but it is more efficient than the one observed for the p. Cys317Tyr mutant. N=6, 4, and 2 for WT, p. Thr62Pro and p. Cys317Tyr, respectively; ^*^p<0.05, ^***^p<0.001, ^****^p<0.0001 using one way ANOVA, followed by Bonferroni’s multiple comparison test post hoc test. (C) Immunofluorescence analysis of HEK293 cells 10 h after transfection with the indicated UMOD isoform. UMOD is seen in red (stained with HA) and the plasma membrane in green (stained with streptavidin-FITC after biotinylation) in the merged picture. Bar=10 µm. The graph reports the Mander’s 2 coefficient as a read-out for UMOD at the plasma membrane. ^**^p<0.01, ^***^p<0.001, ^****^p<0.0001 using one way ANOVA, followed by Bonferroni’s multiple comparison test post hoc test. (D) GRP78 (*HSPA5*) and spliced XBP1 expression assessed by real-time quantitative PCR in HEK 293 cells expressing the indicated UMOD isoform in basal condition (top panels) or after 12 hours treatment with a low dose of tunicamycin (bottom panels). Expression is normalised to *HPRT1*. ^*^p<0.05, ^**^p<0.01 ^****^p<0.0001 using one way ANOVA, followed by Bonferroni’s multiple comparison test post hoc test (n = 3-5 independent experiments). (E) Immunofluoresecence analysis of unpermeabilized MDCK cells stably showing the indicated UMOD isoform at the plasma membrane. All the analyzed variants form polymers on the plasma membrane while the pathogenic ADTKD mutant p.Cys150Ser forms aggregates. Bar=10 µm. Abbreviations: ER, endoplasmic reticulum; PM, plasma membrane.

Yet, p.Thr62Pro, p.Leu180Val and p.Thr469Met UMOD isoforms are all able to form extracellular polymeric filaments similar to the ones observed for the wild-type protein, contrasting with pathological aggregates observed for p.Cys150Ser in MDCK cells (Figure 2E). These data reveal a unique UMOD isoform p.Thr62Pro, with an intermediate defect in processing from ER to Golgi, contrasting with wild-type and other low-frequency variants p.Leu180Val and p.Thr469Met and with canonical ultrarare ADTKD-*UMOD* mutations. Moreover, under mild stress conditions pThr62Pro induces ER stress at a level that is intermediate between wild-type and a typical ADTKD mutant.

### Modelling of p.Thr62Pro effect on protein folding

UMOD contains 48 conserved cysteine residues which are all engaged in intramolecular disulphide bonds (21) (Figure S3). There is a major enrichment of cysteine substitutions in patients with ADTKD-*UMOD* (∼55% of all missense variants) (11), such that most cysteine positions in UMOD (35/48, 73%) have been substituted in patients with ADTKD-*UMOD* (Table S3) vs. only 10% (5/48) in gnomAD. In contrast, all the other amino acids, apart from tryptophan, show more genetic variability in gnomAD compared to the ADTKD population, supporting that cysteine residues in UMOD are intolerant to genetic variation (Figure S4A). In fact, Thr62 is directly adjacent to Cys63 that is predicted to form a disulphide bridge with Cys52 (from Uniprot, PROSITE-ProRule:PRU00076) (22) (Figure 3A, top and middle), and substitutions of Cys52 and Cys63 are identified in multiple ADTKD-*UMOD* families (Table S3). We therefore postulated that p.Thr62Pro might affect UMOD maturation by impacting on neighbouring disulphide bridge formation. Proline has a cyclic pyrrolidine side chain determining high conformational rigidity and potential high impact on protein stability (23).

**Figure 3.**
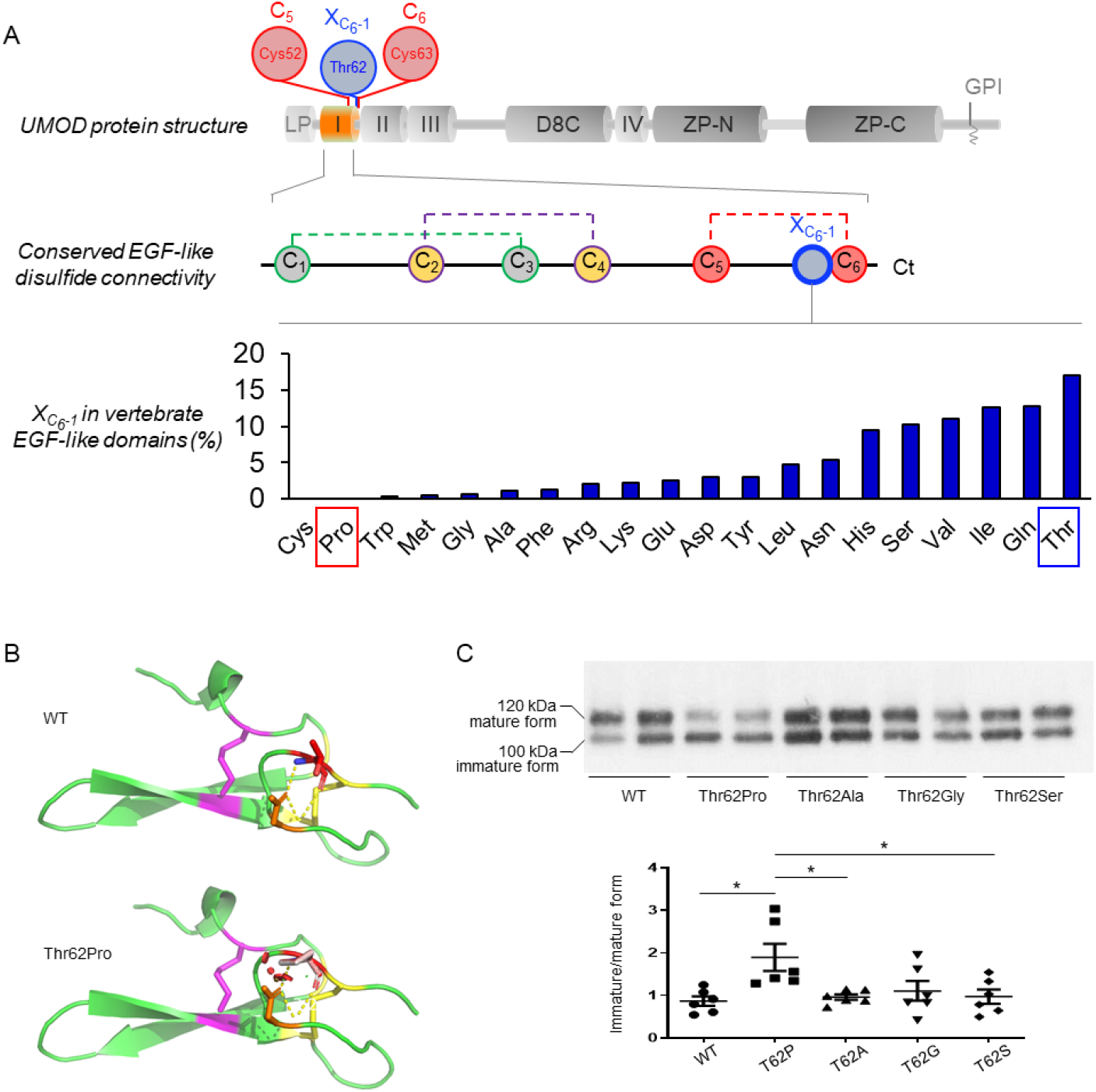
Modelling of p.Thr62Pro impact on protein structure. (A) (Top) Schematic representation of UMOD domain structure: LP, leader peptide; I-IV, EGF-like domains; D8C, domain with 8 cysteines; ZP, bipartite Zona Pellucida domain; GPI, GPI anchoring site. The EGF-like domain 1 is shown in orange; the position of Cys52 and Cys63, predicted to form a disulphide bond, and of neighboring Thr62, is shown. (Middle) Schematic representation of the EGF-like domain showing the conserved disulphide bond connectivity (C1-C3; C2-C4; C5-C6) (data from PROSITE documentation PDOC00021). The position corresponding to Thr62 in UMOD is indicated in blue (1 amino acid before the sixth cysteine, X_C6-1_). (Bottom) Vertebrate EGF-like domain sequences from Pfam family PF12947 (8,511 sequences) were aligned and the frequency of residues at the position preceding C6 (XC6-1) was calculated. Pro and Cys are the only amino acids that are never found, strongly suggesting a damaging effect of Pro at this position. Note that Thr is the most frequently encountered amino acid at position X_C6-1_. (B) Modelling of the UMOD p.Thr62Pro isoform. Polar contacts are indicated by yellow dashed line. Clashes in the structure are represented by red dots and are visible only in the p.Thr62Pro isoform. The figure was made with PyMOL (Schrödinger LLC). (C) Western blot analysis for UMOD in cell lysates from HEK293 cells transiently transfected with the indicated UMOD isoforms. The immature and mature forms of UMOD proteins are indicated on the left. The ratios of the intensities of immature/mature forms are indicated below. Note that only the proline substitution increases the ratio of immature/mature form indicating defective trafficking. N=6 independent experiments; ^*^p<0.05 using one way ANOVA, followed by Bonferroni’s multiple comparison test post hoc test.

Thus, missense variants replacing an amino acid with a proline are poorly tolerated when adjacent to a cysteine bond (24). Four different cysteine-adjacent positions are substituted to proline in ADTKD-*UMOD* families, more than for any other amino acid in UMOD (Figure S4B). In line, when aligning >8,000 vertebrate EGF-like domain sequences, we observed that proline is never present at the position corresponding to Thr62, i.e., preceding the sixth cysteine of the domain (X_C6-1_), while all the other amino acids except cysteine can be found (Figure 3A), further supporting a damaging effect of proline at this position. To analyse the effect of the substitution, we constructed a model of the first EGF of UMOD (aa 28-65) using the software Phyre2 and introduced the threonine to proline change in Pymol. The presence of structural clashes is detected when proline is inserted (Figure 3B). In contrast, non-proline substitutions at Thr62 in UMOD appear tolerated (Figure S4C & S5). These results were confirmed by using the prediction programs Missense3D (25) and SDM (Figure S4C) (26), which identified proline as the only substitution affecting protein structure and as the most destabilizing. Furthermore, only Thr62Pro, but not Thr62Ala, nor Thr62Gly or Thr62Ser, led to a trafficking defect in HEK293 cells (Figure 3C). Collectively, these results strongly suggest a unique effect of p.Thr62Pro on protein maturation, likely via an impact on the conserved neighbouring disulphide bridge within the canonical EGF-like domain.

### High genetic load of UMOD p.Thr62Pro variant in familial tubulointerstitial CKD clusters

The *UMOD* p.Thr62Pro variant was initially detected in proband III.1 of family CH1, presenting with CKD, hyperuricemia with gout, small multicystic kidneys, normal urinalysis and negative for other ADTKD genes (Figure S6 & Table S4). However, the p.Thr62Pro variant did not fully segregate with disease in this family, being detected in apparently unaffected relatives (Figure S6). The variant was also reported in a German family (labelled here as GE1) as variant of unknown significance (VUS) segregating with ADTKD. Kidney biopsy revealed interstitial fibrosis with tubular atrophy (27). Carriers of p.Thr62Pro were identified in 9 additional ADTKD families (Figure S6) and in 5 unrelated probands with kidney disease (i.e., CKD, gout, kidney cysts) in the UK 100,000 Genomes Project (Figure S6 & Table S4). No alternative variants were detected in known ADTKD genes (Table S4) and after whole genome sequencing (WGS) in the families from the 100,000 Genomes Project. Segregation analysis confirmed incomplete penetrance for the p.Thr62Pro variant, also detected in unaffected carriers (Figure S6: CH1: II.2, III.2; UK2: IV3 and GEL4: I.1). Incomplete penetrance (or late onset disease) is also evident in the 100,000 Genomes Project where p.Thr62Pro was detected in 52 unaffected relatives and 57 rare disease patients *without* a primary kidney diagnosis and with an average age of 41.5 years (Table S5 & Table S6). Thus, the p.Thr62Pro variant is detected in a significant number of families with tubulointerstitial CKD of unknown origin and evidence for dominant inheritance. Combining the ADTKD-*UMOD* families reported thus far (Table S3) and including the families from this study, p.Thr62Pro is detected in ∼7% (16/243) of them. This corresponds to an AF for p.Thr62Pro of ∼0.033 (16/486) among ADTKD family index patients, ∼100x increased compared to the AF for p.Thr62Pro in gnomAD (∼0.00035).

### Milder phenotype in p.Thr62Pro carriers with kidney disease

We next tested whether UMOD p.Thr62Pro was associated with an intermediate phenotype, consistent with its biological effect. Analysis of 32 p.Thr62Pro carriers that showed signs of kidney disease (eGFR<60mL/min/1.73m^2^, excluding young or unaffected carriers; Table S4) compared to ADTKD-*UMOD* patients carrying canonical *UMOD* mutations indicated that p.Thr62Pro was associated with a significantly longer kidney survival (median age of kidney failure: 69 years vs. 54 years, log rank P=0.001) (Figure 4A), with a mean age at kidney failure of 57.4±15.7yr vs. 47.5±12.4yr, respectively (P=0.002) (Figure 4B). Markedly decreased levels of UMOD in the urine (uUMOD) are a hallmark of ADTKD-*UMOD*, reflecting mutant UMOD retention in the kidney (11). Compared to uUMOD levels in control individuals (17±10mg/g creatinine) and ADTKD-*UMOD* patients (2±2mg/g creatinine), intermediate levels were observed in p.Thr62Pro carriers (9±5mg/g creatinine) (Kruskall-Wallis P<0.0001). The latter never showed the very low uUMOD concentrations (<1µg/mL) often detected in ADTKD-*UMOD* samples (Figure 4C & Table S7). Comparative immunofluorescence studies in human kidney samples revealed increased intracellular UMOD staining with somewhat preserved apical enhancement in a p.Thr62Pro carrier, as compared with strongly predominant apical staining in normal kidney tissue (from a tumour nephrectomy sample) and exclusive intracellular staining in a patient harbouring a canonical mutation (p.Arg185Ser) for ADTKD-*UMOD*. Of note, a discrete upregulation of ER-stress marker GRP78 was noted in UMOD-positive tubules in the p.Thr62Pro sample, much weaker than that observed in canonical ADTKD-*UMOD* (Figure 4D). These data indicate that the fraction of *UMOD* p.Thr62Pro carriers who presents with CKD shows a milder kidney disease, with progression to kidney failure delayed by ∼15 years compared to classical ADTKD-*UMOD*, paralleled by intermediate levels of uUMOD reduction reflecting a milder trafficking defect and intracellular UMOD accumulation.

**Figure 4.**
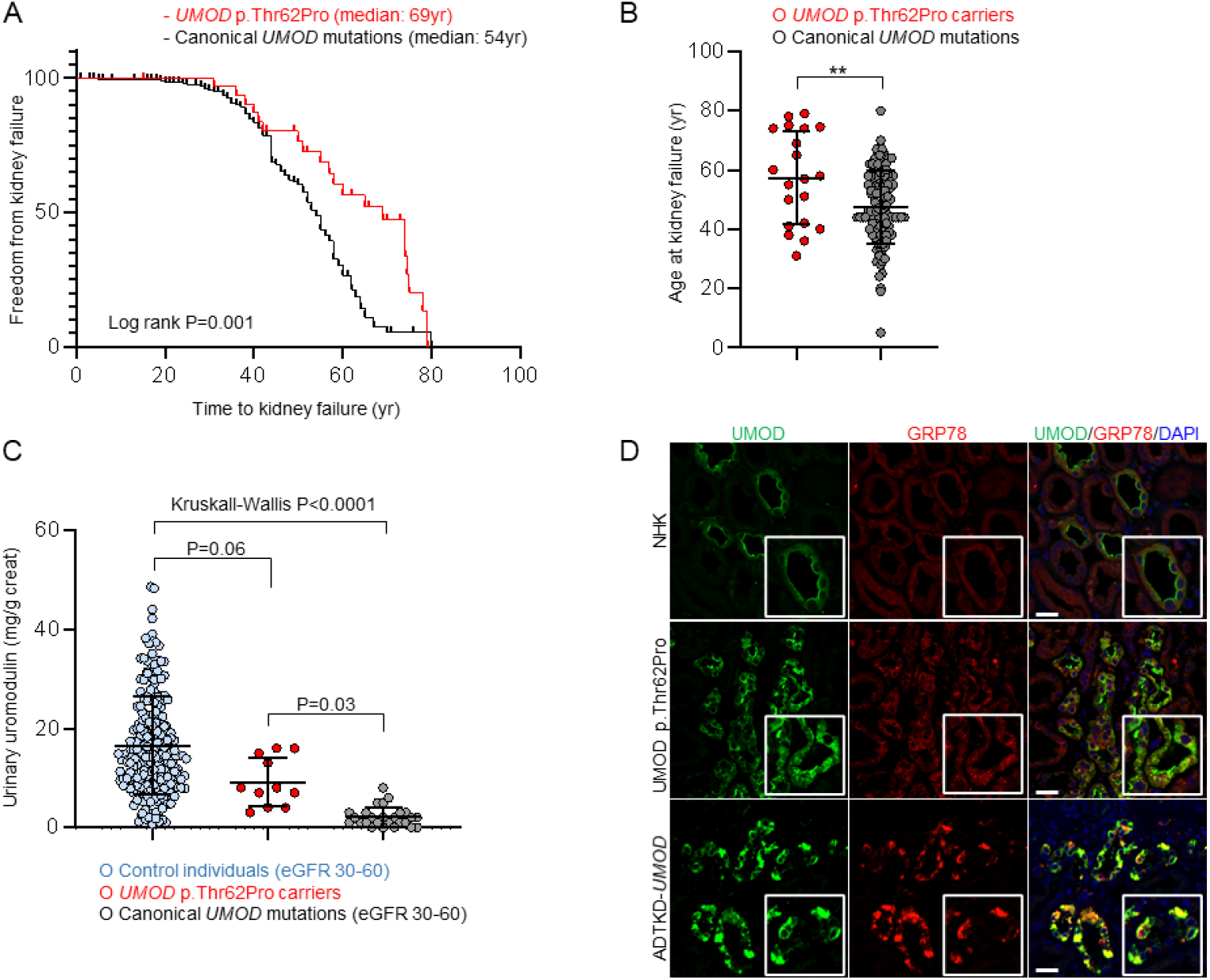
Mild kidney disease phenotype and intermediate UMOD trafficking defect in p.Thr62Pro carriers with CKD. (A) Kaplan-Meier curve of renal survival in p.Thr62Pro carriers with CKD and in ADTKD-*UMOD* patients from the International ADTKD Cohort. Only patients with known p.Thr62Pro status and signs of kidney disease (GFR<60mL/min/1.73m^2^) have been included in the analysis (n=32). Median age at kidney failure was 69 years in *UMOD* p.Thr62Pro carriers and 54 years in ADTKD-*UMOD* patients with canonical *UMOD* mutations. (B) Scatter plots for age at kidney failure for p.Thr62Pro carriers with kidney disease and ADTKD-*UMOD* patients with canonical *UMOD* mutations. Bars indicate means±SD. ^**^p<0.01 using unpaired t test. (C) Urinary UMOD levels normalized to urinary creatinine are depicted for control individuals from the Cohorte Lausannaise (CoLaus) matched for eGFR (30-60mL/min/1.73m^2^), *UMOD* p.Thr62Pro carriers and ADTKD-*UMOD* patients with canonical *UMOD* mutations matched for eGFR (30-60mL/min/1.73m2). Outlier removal performed using ROUT (Q=1%). Graph depicts individual values and bars indicate mean±SD. Kruskall-Wallis test was used with Dunn’s multiple comparisons test. (D) Immunofluorescence staining for UMOD (green) and glucose-regulated protein 78 (GRP78; red) in normal human kidney (NHK, from a tumor nephrectomy), *UMOD* p.Thr62Pro human kidney biopsy and kidney tissue from an ADTKD-*UMOD* patient with a canonical *UMOD* mutation (p.Arg185Ser). Bars=25 μm. DAPI, 4′,6-diamidino-2-phenylindole.

### UMOD p.Thr62Pro is enriched in patients with kidney failure

To investigate whether intermediate-effect *UMOD* variants contribute to risk of CKD, we performed a single variant load analysis in the Genomics England 100,000 Genomes project. Among the top 14 low-frequency (GnomAD AF>10^−3^) or rare (GnomAD 10^−3^>AF>10^− 4^) *UMOD* missense variants (Table S1), pThr62Pro is the only variant with higher prevalence in the three tested modalities 1. 100,000 Genomes kidney disease categories, 2. Human Phenotype Ontology (HPO) term CKD and 3. HPO term CKD stage 5 compared to gnomAD and the 100,000 Genomes control population (Figure S7). Next, we selected 1351 cases (958 probands) in the 100,000 Genomes project with stage 5 CKD, treated by kidney transplantation or dialysis, using the appropriate ICD10 and HPO terms (ICD10: N185, Z940, Z491, Y841 and/or HPO HP:0003774) as well as 87,567 controls (32,371 probands) in which none of these diagnoses were present and tested for the prevalence of p.Thr62Pro (Table S6). A significant enrichment for p.Thr62Pro was detected when considering the whole cohort (OR 2.80, P=0.01) and when considering only probands (OR 3.99, P=0.006) (Figure 5A). As the percentage of individuals with white ethnicity (as provided by Genomics England) was similar between cases and controls, it is unlikely that stratification based on genetic ancestry was driving this association with p.Thr62Pro, which is enriched in Europeans (Table S6). Similar results were observed in ancestry-matched individuals from the UK biobank, where UMOD p.Thr62Pro was enriched in probands with CKD stage 5 and probands with kidney transplantation (OR 4.12, P=0.038 & OR 6.84, P=0.010) (Figure 5B). In both databases, no significant enrichment for p.Thr62Pro was detected for milder stages of CKD. *UMOD* variants p.Leu180Val and p.Thr469Met were not significantly enriched in probands with kidney disease (Figure S8 & S9).

**Figure 5.**
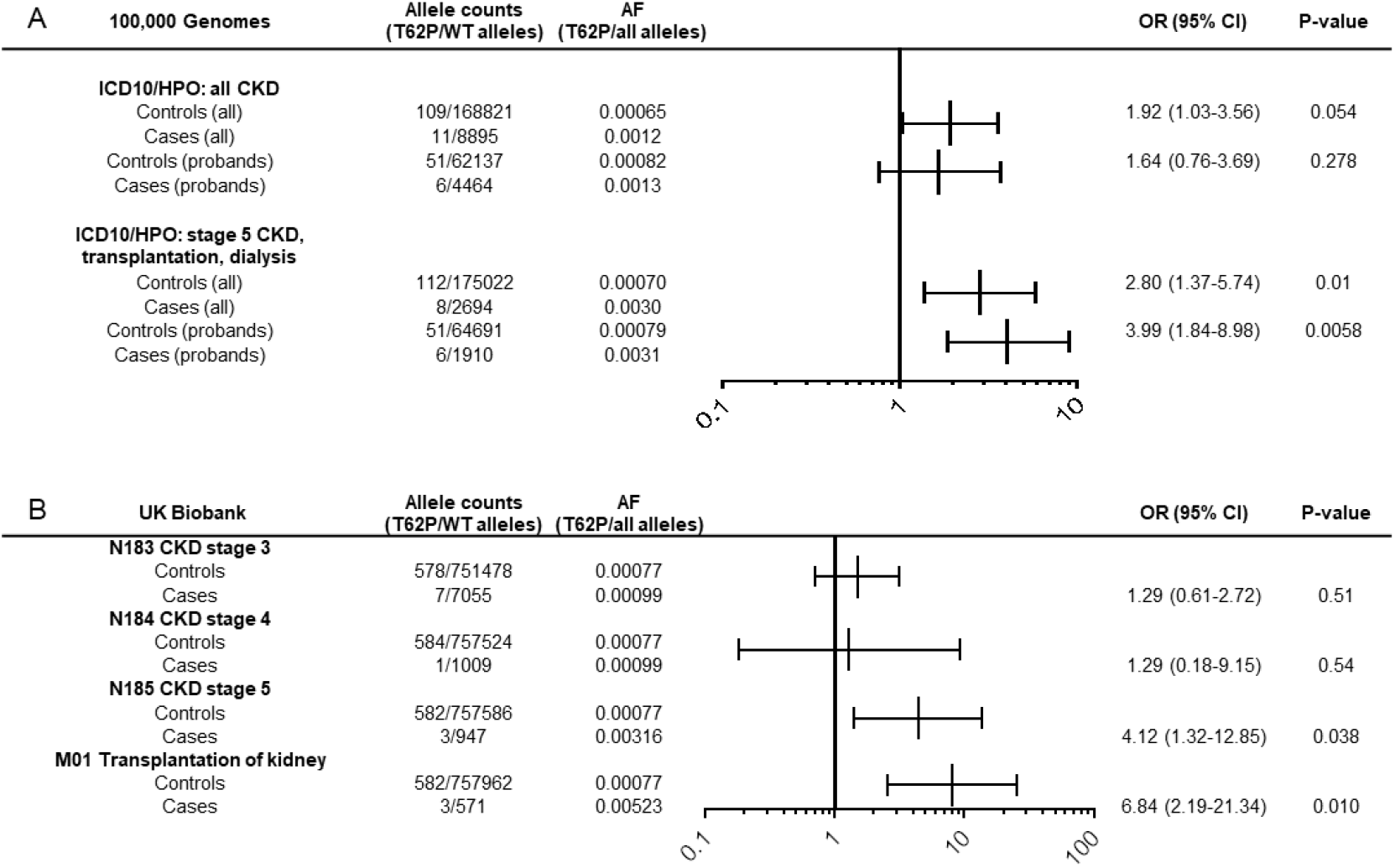
UMOD p.Thr62Pro confers risk for kidney disease in UK cohorts. (A) Prevalence of p.Thr62Pro alleles in all individuals and in probands from the Genomics England 100,000 Genomes project with diagnoses of CKD (ICD10: N18 and/or HPO HP0012622 diagnoses including mapped and descendant concepts) or kidney failure (ICD10: N185, Z940, Z491, Y841 and/or HPO HP:0003774 diagnoses including mapped and descendant concepts) and in all control individuals or family probands in which these diagnoses are absent. P values computed using Fisher’s exact test. (B) Prevalence of p.Thr62Pro alleles in unrelated white British controls and individuals with indicated kidney phenotypes in the UK biobank SNP array data. P value computed using Fisher’s exact test.

## Discussion

The *UMOD* gene is of particular interest because it encodes the most abundant protein excreted in the normal urine and is associated with an autosomal dominant kidney disease (ADTKD-*UMOD*) and is the top GWAS signal for eGFR and CKD in the general population (4). By intersecting missense variants associated with ADTKD-*UMOD* or present in gnomAD, we detected a unique *UMOD* variant p.Thr62Pro with intermediate-size biological effects, associated with kidney failure and enriched in CKD families with a milder clinical course compared to ADTKD-*UMOD*. The identification of an intermediate-size effect variant completes the spectrum of *UMOD*-associated kidney diseases and provides novel insights into the genetic architecture of CKD.

Using large-scale reference datasets, we confirm that *UMOD* variants associated with ADTKD are generally ultrarare and absent from gnomAD, as expected for *bona fide* high-effect variants causing a dominant Mendelian disease (11). The 8 *UMOD* variants detected both in ADTKD patients and gnomAD showed a prevalence of ∼1/160 in gnomAD, similar to the prevalence (∼1/165) of purported pathogenic variants in *PKD1* reported in ExAC (28). These observations could be due to either annotation errors or altered penetrance and expressivity of purported pathogenic variants in control datasets. The MAF of p.Thr62Pro in the general population (between 0.035% and 0.05%) and the fact that it is among the most frequent *UMOD* missense variants listed in gnomAD is not compatible with a high-effect size variant for ADTKD-*UMOD*, whose prevalence is estimated between 1:50,000 and 1:100,000 (10). However, p.Thr62Pro is enriched in kidney disease populations, with a MAF of ∼0.4% among UK patients with kidney failure and up to 3% among ADTKD family index patients. These epidemiological data are compatible with UMOD p.Thr62Pro occupying an intermediate position in terms of allelic frequencies between common and ultrarare variants and also compatible with an intermediate effect size.

What could be the basis for the increased risk of CKD associated with the *UMOD* p.Thr62Pro variant? While ADTKD-*UMOD* is caused by gain-of-toxic-function mutations altering ER homeostasis (10, 29, 30), the risk of CKD associated with common GWAS *UMOD* variants is thought to be related to increased UMOD expression which may, over time, stress the ER protein folding capacity and lead to tubulointerstitial damage (5). Our data suggest that the same cellular pathways, involving UMOD ER homeostasis and maturation, may sustain the CKD risk conferred by the p.Thr62Pro *UMOD* variant. The variant shows intermediate trafficking defect and induction of ER stress between wild-type and canonical ADTKD mutant isoforms. Interestingly, induction of ER stress is only observed under mild tunicamycin treatment that affects ER homeostasis, an experimental setting that could mimic the reduction of chaperoning systems in aging cells (31). Consistently, patients with the p.Thr62Pro variant show intermediate urinary UMOD levels, paralleled by mild intracellular accumulation of UMOD and ER stress. Such significant but mild biological effects match the intermediate clinical effect sizes for p.Thr62Pro, as evidenced by significantly longer kidney survival and older age at kidney failure compared to those observed for the canonical ADTKD-*UMOD* mutations. Furthermore, the *in vitro* data suggest that additional stressors may be required to induce cell toxicity, possibly reflected by the incomplete penetrance and later onset of disease in pThr62Pro carriers. These findings also corroborate the recent evidence that, in ADTKD-*UMOD*, the severity of the mutant UMOD trafficking defect correlates with disease severity (20). Further *in vitro* and *in silico* evidence substantiate the specific effect of the proline substitution on UMOD folding and ER exit, possibly by impacting on the neighbouring disulphide bond, in line with mutagenesis datasets showing missense proline insertions being associated with highest predicted damage (32). Whether and how p.Thr62Pro could have additional, deleterious effects on the function of UMOD remains to be investigated.

While the majority of UMOD p.Thr62Pro carriers do not develop overt kidney disease, the variant appears to segregate with CKD in some families. The basis for such heterogeneity remains speculative. No other known pathogenic variants were detected in these families, 5 of which have undergone WGS. One could speculate that other genetic factors, possibly in addition to environmental factors, could modulate the threshold for disease expression and CKD (15, 33). Such modifiers could involve the ER-proteostasis pathway, in line with induction of ER stress in p.Thr62Pro expressing cells upon mild tunicamycin treatment, or genes that determine the inflammatory response (10). Our data indicate that CKD progression is delayed in p.Thr62Pro carriers and disease might be subclinical in some younger adults. In line, we saw that p.Thr62Pro carriers that were labelled with CKD in the 100,000 Genomes dataset were notably older than carriers in which no CKD was described (Table S6). The true life-long risk for developing CKD in ageing p.Thr62Pro carriers thus remains to be determined.

Thus far, only a few examples of genes showcasing a spectrum of effect sizes have been documented. Reported pathogenic variants in the prion protein gene *PRNP*, causing autosomal dominant prion disease due to gain-of-function prion protein misfolding, are at least 30× more common in the population than expected based on disease incidence (34). The authors suggested that some of these variants confer intermediate amounts of lifetime risk rather than being fully penetrant. About 1% of individuals from the general population carry *bona fide* pathogenic variants in Mendelian diabetes mellitus genes, yet remain euglycemic through middle age (35). Paradigmatically, *HNF1A* has been implicated in a spectrum of phenotypes ranging from multifactorial polygenic diabetes risk to autosomal dominant early-onset diabetes (36). Whole-exome sequencing of Latino populations has revealed a single low-frequency missense variant (p.Glu508Lys) in *HNF1A* associated with an intermediate functional effect and a 5-fold increase in type 2 diabetes prevalence. Of note, only 1 variant in *HNF1A* and 3 variants in *PRNP* have been associated with intermediate effect sizes, compatible with the single variant described here (34, 37). The identification of additional ultrarare variants with intermediate-effect sizes contributing to diverse human phenotypes may require even better powered studies.

The identification of an *UMOD* variant with incomplete penetrance has implications for genetic testing and counselling. Indeed, the implementation of NGS in routine care means that more VUS in *UMOD* will be identified, requiring careful characterization and mapping into the disease spectrum. Previous analyses reported an absence of positive family history in ∼15% of cases as well as incomplete penetrance in ADTKD-*UMOD* (38–40), suggesting the possibility of further, atypical *UMOD* variants with intermediate effect sizes. Several atypical *UMOD* variants were identified here (Table 1) but, in contrast to p.Thr62Pro, with very low population frequencies (MAF ∼10^−5^). The fact that numerous ultrarare and predicted pathogenic *UMOD* variants are present in gnomAD is noteworthy (Table S2). If the 45 individuals harbouring such ultrarare variants with a REVEL score >0.75 (corresponding to a specificity of ∼0.95 for pathogenic variants (19), and excluding the probable benign variant p.Gly131Asp) would develop mild or late-onset CKD, then the prevalence of ADTKD-*UMOD* would rise close to 1 in 3,000 (45 carriers in ∼140,000 gnomAD subjects).

This study is the first systematic investigation of intermediate-size effect of *UMOD* variants, based on analysis of large databases, detailed characterization of the effect *in vitro*, by *in silico* modelling and in patient samples, and validation in large patient and population-based cohorts. Limitations of the study include using previously reported ADTKD-*UMOD* variants, rather than re-sequencing patients, thereby limiting the discovery stage. Most of the patient and control dataset is of European Ancestry, thus neglecting potential variants operating in other backgrounds. For instance, the p.Leu180Val *UMOD* variant, ultrarare in UK databases, is principally detected in Africans. Approximately 1 in 1,000 people of European ancestry carry p.Thr62Pro: based on our findings, these individuals have a 4-fold increase in risk for kidney failure. Longitudinal follow-up studies are required to determine the clinical significance of the p.Thr62Pro carrier status in the general population and whether UMOD p.Thr62Pro could also act as a modifier in other types of kidney diseases.

In conclusion, we identified a single rare missense variant (p.Thr62Pro) in *UMOD*, characterized by an intermediate biological effect *in vitro* and *in vivo*, associated with a milder course of ADTKD and conferring an increased risk for kidney failure in UK cohorts. These findings extend the understanding of *UMOD*-associated kidney diseases and of the genetic architecture of CKD.

## Materials and Methods

A detailed description of the materials and methods (patient and population cohorts, *in vitro* and *in silico* analyses, statistics) is presented in the supplementary information. Informed and written consent was obtained from all participants and individual-level data was de-identified. All methods were carried out in accordance with approved guidelines and the Declaration of Helsinki. The 100,000 Genomes research and clinical project model and its informed consent process has been approved by the United Kingdom National Research Ethics Service Research Ethics Committee for East of England – Cambridge South Research Ethics Committee (https://www.genomicsengland.co.uk/about-genomics-england/the-100000-genomes-project/)(Ref 14/EE/1112). Ethics approval for the UK Biobank study was obtained from the United Kingdom North West Centre for Research Ethics Committee (approval number 11/NW/0382) (https://www.ukbiobank.ac.uk/). The German Chronic Kidney Disease Cohort was approved by the ethics commission of the Friedrich-Alexander-Universität Erlangen-Nürnberg, Germany (Nr 3831) and by the ethics committees of all participating institutions and registered in the national registry for clinical studies (Deutsches Register Klinischer Studien 00003971) (https://www.gckd.de/). The ADTKD cohort study was approved by the institutional review board of the Wake Forest School of Medicine, North Carolina, USA (Wake Forest University Health Sciences IRB00000352 “Characteristics of Individuals with Inherited Kidney Disease”), the Institutional review board of the Université Catholique de Louvain (UCL) Medical School and Saint Luc University Hospital, Belgium (2011/04MAI/184) and the European Community’s Seventh Framework Program “European Consortium for High-Throughput Research in Rare Kidney Diseases (EURenOmics) Ethics Advisory Board. Further institutions involved in human participant research received local IRB approval as detailed in <u>Table S8</u>.

## Data Availability

The data that support the findings of this study are available from the article and the supplementary files, through The Human Gene Mutation Database at http://www.hgmd.cf.ac.uk/ac/index.php and PubMed at https://pubmed.ncbi.nlm.nih.gov. Data presented here were derived from the following resources available in the public domain: The Genome Aggregation Database v2.1.1 (https://gnomad.broadinstitute.org/) and Ensembl (release 100): (https://www.ensembl.org/index.html).

## Data Availability

The data that support the findings of this study are available from the article and the supplementary files, through The Human Gene Mutation Database at http://www.hgmd.cf.ac.uk/ac/index.php and PubMed^®^ at https://pubmed.ncbi.nlm.nih.gov. Data presented here were derived from the following resources available in the public domain: The Genome Aggregation Database v2.1.1 (https://gnomad.broadinstitute.org/) and Ensembl (release 100): (https://www.ensembl.org/index.html). The International ADTKD Cohort as well as the UMOD *in vitro* scoring have been published (11, 20).

## Acknowledgments

We thank all participating patients and families. The Cohorte Lausannoise is acknowledged for providing reference urine samples and eGFR information. We are grateful to the Broad Institute of MIT and Harvard, the First Faculty of Medicine, Charles University (LM2018132), and the Friedrich-Alexander-Universität Erlangen-Nürnberg in Germany for providing *MUC1* testing. The authors are grateful to Anne Kipp for her help in generating preliminary results and data interpretation and to Valeria Berno from San Raffaele Advanced Light and Electron Microscopy BioImaging Center for helpful discussions. EO is supported by an Early Postdoc Mobility-Stipendium of the Swiss National Science Foundation (P2ZHP3_195181) and a Kidney Research UK grant Paed_RP_001_20180925, the Fonds National de la Recherche Luxembourg (grant 6903109) and the University Research Priority Program “Integrative Human Physiology, ZIHP” of the University of Zurich. MW is supported by Deutsche Forschungsgemeinschaft (DFG) – Projektnummer 387509280 – SFB 1350, TP C4. The work of AK was supported by German Research Foundation (DFG) grant KO 3598/5-1, DFG - Project-ID 431984000 - SFB 1453. ADK was supported by a grant from the University of Zurich (Forschungskredit FK-14-035). EAEE is funded by the Royal College of Surgeons in Ireland Blackrock Clinic StAR MD. OD is supported by the European Union’s Horizon 2020 research and innovation program under the Marie Skłodowska-Curie grant agreement No 860977, the European Reference Network for Rare Kidney Diseases (project ° 739532), the Swiss National Science Foundation’s National Center of Competence in Research Kidney Control of Homeostasis program, the Swiss National Science Foundation (grant 310030-189044, the Gebert-Rüf Foundation for research on ADTKD-*UMOD* and the University Research Priority Program (URPP) ITINERARE at the University of Zurich. KK, SK and AJB were supported by grant NU21-07-00033 from the Ministry of Health of the Czech Republic. LR is supported by the Italian Ministry of Health (RF 2010 2319394 and RF 2016 02362623). JAS is supported by Kidney Research UK and the Northern Counties Kidney Research Fund. This research was made possible through access to the data and findings generated by the 100,000 Genomes Project. The 100,000 Genomes Project is managed by Genomics England Limited (a wholly owned company of the Department of Health and Social Care). The 100,000 Genomes Project is funded by the National Institute for Health Research and NHS England. The Wellcome Trust, Cancer Research UK and the Medical Research Council have also funded research infrastructure. The 100,000 Genomes Project uses data provided by patients and collected by the National Health Service as part of their care and support. See supplemental information for consortium details. This research has been conducted using data from UK Biobank (project ID 43879), a major biomedical database with open access to researchers (www.ukbiobank.ac.uk). UK Biobank is supported by its founding funders the Wellcome Trust and UK Medical Research Council, as well as the Department of Health, Scottish Government, the Northwest Regional Development Agency, British Heart Foundation and Cancer Research UK. The organisation has over 150 dedicated staff members based in multiple locations across the UK. The GCKD study was funded by the German Ministry of Research and Education (Bundesminsterium für Bildung und Forschung, BMBF, grant number 01ER0804, K.U.E.); by the Foundation KfH Stiftung Präventivmedizin and by the Deutsche Forschungsgemeinschaft (DFG, German Research Foundation) – Projektnummer 246781735 – SFB 1140; and by grants provided by Bayer, Fresenius Medical Care and Amgen. Genotyping was supported by Bayer Pharma AG. uUMOD measurements in GCKD were supported by the Swiss National Centre of Competence in Research Kidney Control of Homeostasis program and the Swiss National Science Foundation grant 310030_189044.

## Notes

**Competing Interest Statement:** The authors declare no potential conflict of interest relevant to this article.

### Competing Interest Statement

The authors have declared no competing interest.

### Author Declarations

Informed and written consent was obtained from all participants and individual-level data was de-identified. All methods were carried out in accordance with approved guidelines and the Declaration of Helsinki. The 100,000 Genomes research and clinical project model and its informed consent process has been approved by the United Kingdom National Research Ethics Service Research Ethics Committee for East of England - Cambridge South Research Ethics Committee (https://www.genomicsengland.co.uk/about-genomics-england/the-100000-genomes-project/)(Ref 14/EE/1112). Ethics approval for the UK Biobank study was obtained from the United Kingdom North West Centre for Research Ethics Committee (approval number 11/NW/0382) (https://www.ukbiobank.ac.uk/). The German Chronic Kidney Disease Cohort was approved by the ethics commission of the Friedrich-Alexander-Universitaet Erlangen-Nuernberg, Germany (Nr 3831) and by the ethics committees of all participating institutions and registered in the national registry for clinical studies (Deutsches Register Klinischer Studien 00003971) (https://www.gckd.de/). The ADTKD cohort study was approved by the institutional review board of the Wake Forest School of Medicine, North Carolina, USA (Wake Forest University Health Sciences IRB00000352 "Characteristics of Individuals with Inherited Kidney Disease"), the Institutional review board of the Universite Catholique de Louvain (UCL) Medical School and Saint Luc University Hospital, Belgium (2011/04MAI/184) and the European Community's Seventh Framework Program "European Consortium for High-Throughput Research in Rare Kidney Diseases (EURenOmics)" Ethics Advisory Board. Further institutions involved in human participant research received local IRB approval as detailed in Table S8.

