## Supplementary Information for "An intermediate effect size variant in *UMOD* confers risk for chronic kidney disease"

**This PDF file includes:**

Appendix: Genomics England Research Consortium
Supplementary Material and Methods
Figures S1 to S9
Tables S1 to S8
Supplementary Information References

### Appendix: Genomics England Research Consortium

Ambrose, J. C. [1] ; Arumugam, P.[1] ; Bleda, M. [1] ; Boardman-Pretty, F. [1,2] ; Boustred, C. R. [1] ; Brittain, H.[1] ; Caulfield, M. J.[1,2] ; Chan, G. C. [1] ; Fowler, T. [1] ; Giess A. [1] ; Hamblin, A.[1] ; Henderson, S.[1,2] ; Hubbard, T. J. P. [1] ; Jackson, R. [1] ; Jones, L. J. [1,2] ; Kasperaviciute, D. [1,2] ; Kayikci, M. [1] ; Kousathanas, A. [1] ; Lahnstein, L. [1] ; Leigh, S. E. A. [1] ; Leong, I. U. S. [1] ; Lopez, F. J. [1] ; Maleady-Crowe, F. [1] ; Moutsianas, L. [1,2] ; Mueller, M. [1,2] ; Murugaesu, N. [1] ; Need, A. C. [1,2] ; O'Donovan P. [1] ; Odhams, C. A. [1] ; Patch, C. [1,2] ; Perez-Gil, D. [1] ; Pereira, M. B. [1] ; Pullinger, J. [1] ; Rahim, T. [1] ; Rendon, A. [1] ; Rogers, T. [1] ; Savage, K. [1] ; Sawant, K. [1] ; Scott, R. H. [1] ; Siddiq, A. [1] ; Sieghart, A. [1] ; Smith, S. C. [1] ; Sosinsky, A. [1,2] ; Stuckey, A. [1] ; Tanguy M. [1] ; Thomas, E. R. A. [1,2] ; Thompson, S. R. [1] ; Tucci, A. [1,2] ; Walsh, E. [1] ; Welland, M. J. [1] ; Williams, E. [1] ; Witkowska, K. [1,2] ; Wood, S. M. [1,2].

[1] Genomics England, London, UK
[2] William Harvey Research Institute, Queen Mary University of London, London, EC1M 6BQ, UK

### Supplementary Material & Methods

Web Resources

Allele frequency App: http://cardiodb.org/allelefrequencyapp/
cBioPortal MutationMapper: https://www.cbioportal.org/mutation_mapper
ClinVar: https://www.ncbi.nlm.nih.gov/clinvar
Clustal omega: https://www.ebi.ac.uk/Tools/msa/clustalo/
Ensembl: https://www.ensembl.org/index.html
Ensembl VEP: https://www.ensembl.org/info/docs/tools/vep/index.html
FASMA (Formatting and Analysing the Sequences in the Multiple Alignments): http://bioinformatica.isa.cnr.it/FASMA
GnomAD v2.1.1: https://gnomad.broadinstitute.org/
HGMD®: http://www.hgmd.cf.ac.uk/ac/index.php
Image J - Fiji: https://imagej.net/Fiji
Mafft: https://mafft.cbrc.jp/alignment/server/
Missense3D: http://missense3d.bc.ic.ac.uk/missense3d/
Phyre$^2$: http://www.sbg.bio.ic.ac.uk/~phyre2/html/page.cgi?id=index
Pfam: https://pfam.xfam.org/
Progeny pedigree builder: https://pedigree.progenygenetics.com/
Prosite: https://prosite.expasy.org
ProteinPaint: https://pecan.stjude.cloud/proteinpaint (Zhou et al., 2016)
PubMed: https://pubmed.ncbi.nlm.nih.gov/
PyMOL: https://pymol.org/2/
SDM: http://marid.bioc.cam.ac.uk/sdm2/
Varsome®: https://varsome.com/
Online Mendelian Inheritance in Man, OMIM®. McKusick-Nathans Institute of Genetic Medicine, Johns Hopkins University (Baltimore, MD) (Hamosh et al., 2000). World Wide Web URL: https://omim.org/

International ADTKD Cohort

The International ADTKD Cohort consists of patients from the Belgo-Swiss ADTKD registry and the US ADTKD registry and has been previously published (3). The inclusion criteria were those defined by the Kidney Disease: Improving Global Outcomes (KDIGO) consensus (4) including: a family history compatible with autosomal dominant inheritance of CKD with progressive loss of kidney function, bland urinary sediment, absent-to-mild albuminuria and/or proteinuria, normal-sized or small-sized kidneys on ultrasound; and/or (in absence of a positive family history of CKD) a history of early-onset hyperuricemia and/or gout and/or the presence of interstitial fibrosis and/or tubular atrophy on kidney biopsy. Exclusion criteria as previously described (3). Only patients screened for variants in *UMOD* (and *MUC1*) were included in the cohort. Anonymized demographics, clinical and genetic information were recorded in a database. This study was approved by the institutional review board of Wake Forest School of Medicine, the Université Catholique de Louvain (UCL) Medical School, and the European Community's Seventh Framework Programme “European Consortium for High-Throughput Research in Rare Kidney Diseases (EURenOmics)(Table S8). This registry has been supported by the European Reference Network for Rare Kidney Diseases (ERKNet), which is partly co-funded by the European Union within the framework of the Third Health Programme 'ERN-2016-Framework Partnership Agreement 2017-2021. In addition, a cohort of Irish families recruited as part of the Irish Kidney Gene Project (IKGP) and who were subjected to NGS to identify genetic variants underlying kidney disease, were screened for *UMOD* missense variants and three families with *UMOD* p.Thr62Pro were included in this study.

Genetic testing

Informed written consent was obtained from all patients. Genomic DNA was isolated from peripheral blood leukocytes using standard procedures. Direct sequencing of *UMOD* exons was initially performed by Sanger sequencing, as previously described (5). At least exons 3 and 4 were sequenced in all patients enrolled into the International ADTKD Cohort and all 10 coding exons were sequenced in a subset of patients from the Cohort and in all families presented in this manuscript. For genetic testing in the Genomics England 100,000 Genomes project and the UK Biobank, see relevant sections below. *MUC1* genotyping was performed using probe extension assays or *MUC1* VNTR sequencing approaches as previously described (6–8). The tubulopathy gene panel utilises massive parallel sequencing of 37 genes implicated in renal tubulopathies (including *UMOD*, *HNF1B* and *REN*) and has been previously described (9). In addition, direct Sanger sequencing of *REN* exons was performed in some families as previously described (10). The cystic kidney disease panel is a targeted NGS panel including the following genes: *PKD1, PKD2, GANAB, DNAJB11, HNF1B, PKHD1, UMOD, SEC63*, and *PRKCSH,* as previously described (11). In addition, screening for large rearrangements in *HNF1B* was performed using multiplex ligation-dependent probe amplification (MLPA) as previously described (12).

Control population and strategy to identify intermediate-effect *UMOD* variants

The Genome Aggregation Database (GnomAD) v2.1.1 (https://gnomad.broadinstitute.org/) comprises 125,748 exomes and 15,708 genomes sequenced as part of various population genetic studies, totalling 141,456 unrelated individuals from eight major populations (13). Genetic variants are aligned against the GRCh37 genome build and the dataset was released in March 2019. Genetic variants in *UMOD* were filtered for missenses and only those annotated for *UMOD* transcript ENST00000302509.8 were retained. A list of *UMOD* missense variants in gnomAD ("controls") with their allelic frequencies was intersected with *UMOD* variants reported in the International ADTKD Cohort (see above) or in HGMD® ("cases"). Our working hypothesis is based on following assumptions: (i) *UMOD* missense variants in ADTKD cases, in the absence of functional studies or additional genetic arguments (eg. segregation studies), are reported as variant of unknown significance (VUS) and potentially include *bona fide* intermediate-effect variants, (ii) high-effect size variants are in principle too rare for gnomAD (lowest AF in gnomAD: $3.6x10^{-6}$ vs maximum credible population allele frequency for fully penetrant *UMOD* mutations: $1x10^{-7}$ (see below for details) (14), but enriched in ADTKD cases and (iii), low effect variants are not reported as (likely) pathogenic or VUS in ADTKD cases, because of their typical higher allele frequency. In theory, this would lead to a spectrum of very low to high-effect *UMOD* variants enriched at both extremes in control and case groups, respectively. Thus, variants that are shared between controls and cases are candidates for intermediate-effect variants as determined by their obligate intermediate phenotypical effect (non-fully penetrant or milder disease) (Figure 1B).

Maximum credible population allele frequency

The maximum credible population allele frequency and allelic count for pathogenic *UMOD* variants in gnomAD was estimated using the frequency calculator established by Whiffin et al. (14) under the following assumptions: a disease prevalence of 1/50,000; allelic heterogeneity of 1%; genetic heterogeneity of 100%; a penetrance of 100%; a reference population size of 282,000 alleles, and statistical confidence of 0.999.

Genomics England 100,000 Genomes project and variant filtering

All participants in the 100,000 Genomes Project have provided written consent and the 100,000 Genomes project has been approved by the National Research Ethics Service Research Ethics Committee for East of England – Cambridge South Research Ethics Committee (Table S8). Whole genome sequencing (WGS) was performed using the Illumina TruSeq DNA PCR-Free sample preparation kit (Illumina, Inc.) and an Illumina HiSeq 2500 sequencer and reads were aligned to GRCh37 using Isaac Genome Alignment Software (version 01.14; Illumina, Inc.). Variant filtering and annotation was performed as previously described (15). In brief, variants (SNVs, indels) were shortlisted if (i) their MAF in control populations was < 1/1,000 for putative novel causal variants and < 25/1,000 for variants listed as disease-causing in HGMD®, (ii) their predicted impact according to the Variant Effect Predictor (VEP) was "HIGH" or "MODERATE" or if the consequences with respect to the designated transcript included one of "splice_region_variant" or non_coding_transcript_exon_variant" if the variant was in a non-coding gene, and (iii) the variant affected a gene with a known aetiological role in the patient's disease. For each case with prioritised variants, the variant calls, HPO-coded phenotype and the relevant metadata were transferred to Congenica for visualisation in the Sapientia™ web application during multidisciplinary team (MDT) meetings, where each variant was annotated with its likely level of pathogenicity, its contribution to the disease phenotype, and to generate research reports.

UK Biobank

UK Biobank is a large prospective study with over 500,000 participants aged 40–69 years when recruited in 2006–2010 and globally accessible to approved researchers who are undertaking health-related research that's in the public interest (16). Genome-wide genotyping was performed on all UK Biobank participants using the Applied Biosystems UK Biobank Axiom Array. Approximately 850,000 variants, included *UMOD* p.Thr62Pro were directly measured. Furthermore, exome data on ~200,000 individuals have been made available (17). Individuals included in this study (project ID 43879) were those that were determined to have genetic ancestry of "Caucasian" (field ID 22006), were not excluded by genetic relatedness as determined by the ukb_gen_samples_to_remove function in the ukbtools library (18), and had not withdrawn by 20/1/2021. Ethics approval for the UK Biobank study was obtained from the North West Centre for Research Ethics Committee (11/NW/0382) (Table S8).

German Chronic Kidney Disease Cohort

The German Chronic Kidney Disease (GCKD) study is an ongoing prospective multicentre observational cohort and has been previously described (19, 20). In brief, it includes 5,217 patients under regular nephrology care with following inclusion criteria: moderately reduced kidney function defined as eGFR of 30–60 ml/min per 1.73 $m^2$ (stage G3, A1–A3) or an eGFR >60 ml/min per 1.73 $m^2$ in the presence of overt proteinuria (stage G1–G2, A3). Exclusion criteria were non-Caucasian ethnicity, solid organ or bone marrow transplantation, active malignancy within 24 months prior to screening, New York Heart Association Stage IV heart failure, and legal attendance or inability to provide consent. 5,123 participants were genotyped for 2,612,357 markers at the Helmholtz Center Munich using the Illumina Infinium Omni 2.5 Exome-8 microarray (Illumina, GenomeStudio, Genotyping Module Version 1.9.4) and genotype imputation using the 1000 Genomes Phase 3 ALL reference panel was conducted, as previously described (21). All participants provided written informed consent, and the study was approved by the ethics committees of all participating institutions and registered in the national registry for clinical studies (DRKS 00003971) (Table S8).

Analyses of UMOD processing

*Western blot:* HEK293 cells were lysed in octylglucoside lysis buffer (50 mM Tris-HCl, pH 7.4; 150 mM NaCl; 60 mM octyl β-D-glucopyranoside and protease inhibitors cocktail (Merck, Germany)) and analysed by Western blot as described in Schaeffer et al (22), using mouse monoclonal antibody against HA (1:2,000 dilution; Biolegend, San Diego, CA). Quantification was performed using the gel analysis option of ImageJ software (23).

*Pulse-chase experiment:* Pulse chase experiments on HEK293 cells stably expressing UMOD were performed as described in Schaeffer et al (22). UMOD was immunoprecipitated using a sheep anti-UMOD antibody (T0850B, United States Biological, Salem, MA).

*Immunofluorescence on cells:* Immunofluorescence experiments were performed essentially as described in Schaeffer et al (22). UMOD polymers on the surface of MDCK cells stably expressing the indicated UMOD isoform were revealed using an anti-HA antibody (1:500, Biolegend) followed by 1h staining with Alexa-Fluor 594 anti-mouse secondary antibody (1:500; Thermofisher). Images were acquired on an Applied Precision DeltavisionUltra system, using an Olympus 100x 1.4NA oil immersion objective, Z step size of 0.2 µm. Images were deconvolved with Applied Precision's softWorx software (GE Healthcare, Issaquah, WA). To quantify the amount of UMOD on the plasma membrane, immunofluorescence analysis was performed in HEK293 cells 10 hours after transfection of the indicated UMOD isoform. Before staining, cells were incubated 20 min at 4°C with 0.5 mg/mL EZ-Link® Sulfo-NHS-LC-Biotin (Thermofisher). UMOD was revealed with an anti-HA antibody (1:500, Biolegend, San Diego, CA) followed by 1h staining with Alexa-Fluor 594 anti-mouse secondary antibody (1:500; Thermofisher) and the biotinylated membrane with a FITC conjugated streptavidin (1:200, Sigma). All pictures were taken with an UltraVIEW ERS spinning disk confocal microscope (UltraVIEW ERS-Imaging Suite Software, Zeiss 63X/1.4; PerkinElmer Life and Analytical Sciences Boston, MA) and were deconvoluted with Huygens Professional version 19.04 (Scientific Volume

Imaging, The Netherlands). Co-occurrence of UMOD (HA) signal with the one of the membrane (streptavidin) was quantified using the Coloc2 Plugins from ImageJ software (23). For each UMOD isoform between 30 and 50 cells from 3 independent experiments were analysed. We set HA signal as channel 1 and streptavidin signal as channel 2 and the threshold was automatically assigned using the Costes method. Co-occurrence of the two signals was determined using tM1 and tM2 (Manders coefficient above threshold). tM2 can be considered as a readout of UMOD reaching the membrane.

*Immunofluorescence on patient tissue*: Immunodetection of UMOD and GRP78 was performed on 7-μm-thick kidney sections obtained from a biopsy of a UMOD p.Thr62Pro heterozygote with CKD (GE1 III.1), a nephrectomy sample of a patient with canonical ADTKD-*UMOD* (p.Arg185Ser) and normal human kidney (NHK) tissue surrounding a tumour nephrectomy sample. Slides were deparaffinized in xylene and rehydrated in a graded ethanol series. Antigen retrieval was carried out for 10 minutes with citrate buffer (pH 6.0) at 98 °C. After 1h in blocking solution, slides were incubated 1h in room temperature with sheep anti-UMOD primary antibody (1:800; K90071C; Meridian Life Science Inc., Memphis, TN), followed by 1h AlexaFluor488-conjugated donkey anti-sheep (1:400; Invitrogen). The slides were then probed 1h in room temperature with rabbit anti-GRP78 primary antibody (1:400; ab21685; Abcam, Cambridge, UK), followed by 1h incubation with AlexaFluor647-conjugated donkey anti-rabbit antibody (1:400; Invitrogen, Thermo Fisher Scientific). Coverslips were mounted with Prolong gold antifade reagent with 4′,6-diamidino-2-phenylindole (Invitrogen) and analysed under a Leica STELLARIS 5 Confocal Microscope (Leica Camera, Wetzlar, Germany) with a x63/1.4 Plan- Apochromat oil-immersion objective. The use of these samples has been approved by the local Ethical Review Boards.

*Measurements of urinary levels of UMOD:* A validated ELISA method was used to measure urinary uromodulin (uUMOD) levels (second morning urine sample) from patients with ADTKD-*UMOD* and individuals heterozygous for *UMOD* p.Thr62Pro (24). Urinary creatinine was measured using a Synchron DXC800 analyzer (Beckman Coulter, Fullerton, CA). The control samples were obtained from the Cohorte Lausannoise, a population-based study including 6,000 people 35 to 75 years of age from the city of Lausanne, Switzerland (25). Urinary creatinine was used to normalize uUMOD levels as previously described (3). Informed consent was obtained from all participating individuals.

*Measurements of ER stress markers expression:* expression levels of the indicated genes were measured in HEK 293 cells transiently expressing the indicated UMOD isoforms. RNA was extracted with TriFast II (Euroclone, Pero, Italy) following the manufacturer's instructions 72h after transfection. When indicated, cells were treated with tunicamycin (20 ng/ml) for 12 hours before RNA extraction. RNA was retro-transcribed with the iScript gDNA Clear cDNA Synthesis Kit (Bio-Rad, Hercules, CA). Real-time qPCR was performed on the CFX96 Touch instrument (Bio-Rad) using the qPCR Core kit for SYBR® Green I No ROX (Eurogentec, Liège, Belgium) with specific primers for the indicated genes.

| Target gene | Primer Forward (5'>3') | Primer Reverse (5'>3') |
|---|---|---|
| *UMOD* | GCGTACTGCACAGACCCCAGC | GTCATTGAAGCCCGAGCACCG |
| *HSPA5* (Human) | CGCTGAGGCTTATTTGGGAAAG | TGCCGTAGGCTCGTTGATG |
| *XBP1S* (Human) | GAGTCCGCAGCAGGTG | ATACCGCCAGAATCCATGG |
| *HPRT1* (Human) | AGCCCTGGCGTCGTGATTAGTG | TGTGATGGCCTCCCATCTCCTTCA |

*In silico* modelling

*Structural data:* the structure of the first EGF of UMOD was obtained by using the software Phyre2 (26) (submitted sequence aa 25-65, Uniprot P07911-1). We obtained a homology-based model for residues 30-65. The different substitutions were introduced in the obtained PDB formatted model using the mutagenesis wizard in PyMOL, and in Missense 3D and SDM programs.

*EGF-like domain 1 sequence alignment in vertebrates:* The EGF-like domain 1 of human UMOD was analyzed in Pfam, which identified a match with Pfam *EGF_3* (PF12947) family. All vertebrate sequences within PF12947 family were then retrieved (n = 8,622) and aligned with Mafft program. Aligned sequences were manually curated to identify sequences lacking the sixth cysteine ($C_6$). These sequences (n = 222) were scanned in Prosite to verify the presence or absence of the $C_5$-$C_6$ disulphide bond in the EGF-like domain. If this bond was present (n = 111), the missing part of the EGF-like domain sequence was added and realigned. If this bond was reported as absent (n = 111), the sequence was discarded from alignment. We finally retrieved 8,511 aligned sequences that were analysed with FASMA to obtain the number and frequency of the type of amino acid at the position preceding Cysteine 6 ($X_{C6-1}$), corresponding to Thr62 in human UMOD sequence.

Statistics

Categorical variables were compared using the Fisher's exact test. Continuous variables were compared using an unpaired two-tailed *t* test, one way analysis of variance (ANOVA), followed by Bonferroni's multiple comparison post hoc test testing for normally distributed variables or Kruskall-Wallis test with Dunn's multiple comparisons test for non-parametric values. Kaplan-Meier curves were generated to display kidney failure-free survival. Patients who had not reached kidney failure at the end of the study (outcome of interest not occurred during follow-up time) were considered censored individuals. Censoring time was defined as age at last follow-up. A log-rank test was used for comparison of survival curves. Statistical analysis was performed within GraphPad Prism version 9.0.0 for Windows (GraphPad Software, San Diego, California USA, www.graphpad.com). P<0.05 was considered statistically significant.

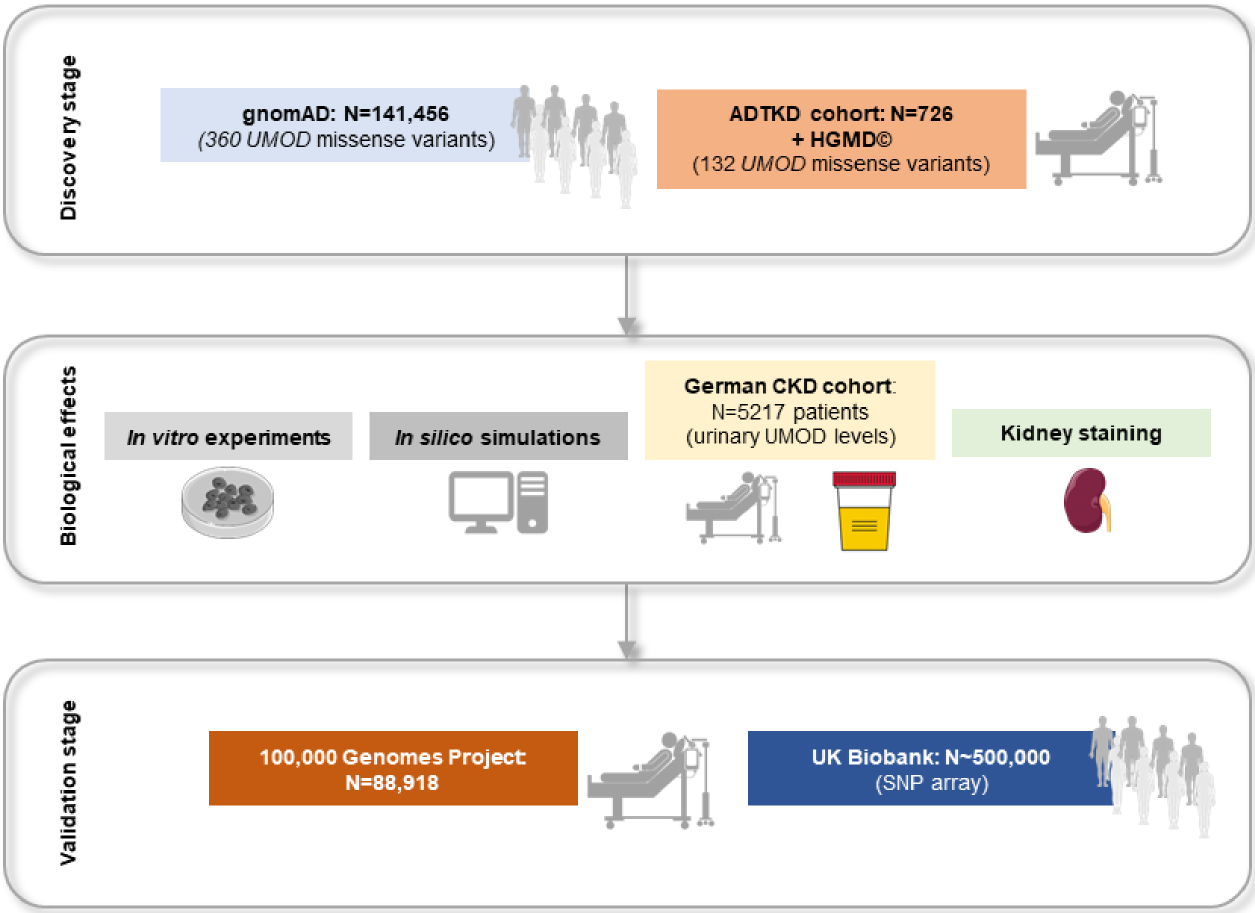


**Fig. S1. Workflow for the identification and validation of intermediate-effect genetic variants in *UMOD*.**

A

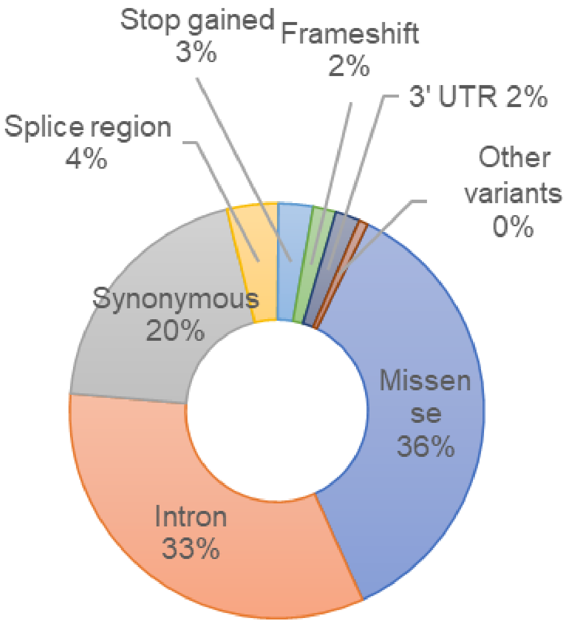


B

| Type of variant | Variant counts |
|---|---|
| Missense | 360 |
| - Cysteine residue affected | 1.4% (5 of 360) |
| - Nonpolar AA affected | 46.1% (166 of 360) |
| - Polar AA affected | 27.8% (100 of 360) |
| - Positively charged AA affected | 13.9% (50 of 360) |
| - Negatively charged AA affected | 12.2% (44 of 360) |
| Intron | 331 |
| Synonymous | 198 |
| Splice region | 40 |
| Stop gained | 27 |
| Frameshift | 18 |
| 3' UTR | 19 |
| Splice donor | 3 |
| Splice acceptor | 3 |
| Inframe deletion | 1 |
| Total | 1000 |

**Fig. S2**. **Landscape of *UMOD* genetic variation in the gnomAD population.** (A) Relative contribution of different classes of genetic alterations in the *UMOD* gene in exome and genome sequencing data from the Genome Aggregation Database (gnomAD v2.1.1) population (141,456 individuals). A total of 1000 variants are reported after removing data related to non-canonical transcripts (81 variants). (B) Quantitative details for *UMOD* variants reported in the gnomAD dataset. Variants denoting non-canonical transcripts have been removed (n=81) and consequences have been checked for consistency with *UMOD* transcript ENST00000302509.8.

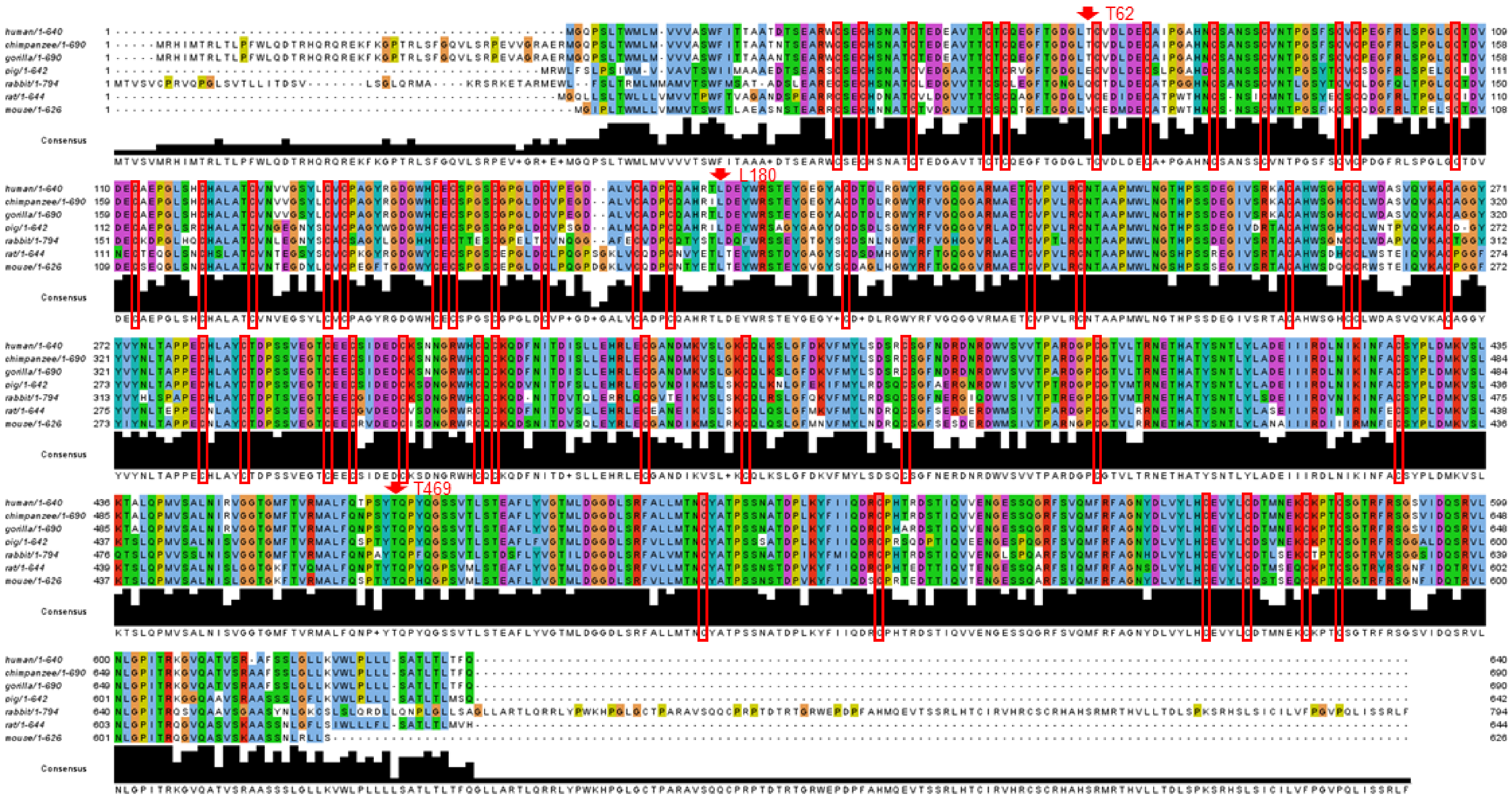


**Fig. S3. UMOD amino acid conservation across mammalian species.** Conservation of UMOD amino acids across indicated mammalian species using Clustal omega. Positions Thr62, Leu180 and Thr469 are indicated by red arrows. Note the 48 cysteine positions (boxed in red), all fully conserved.

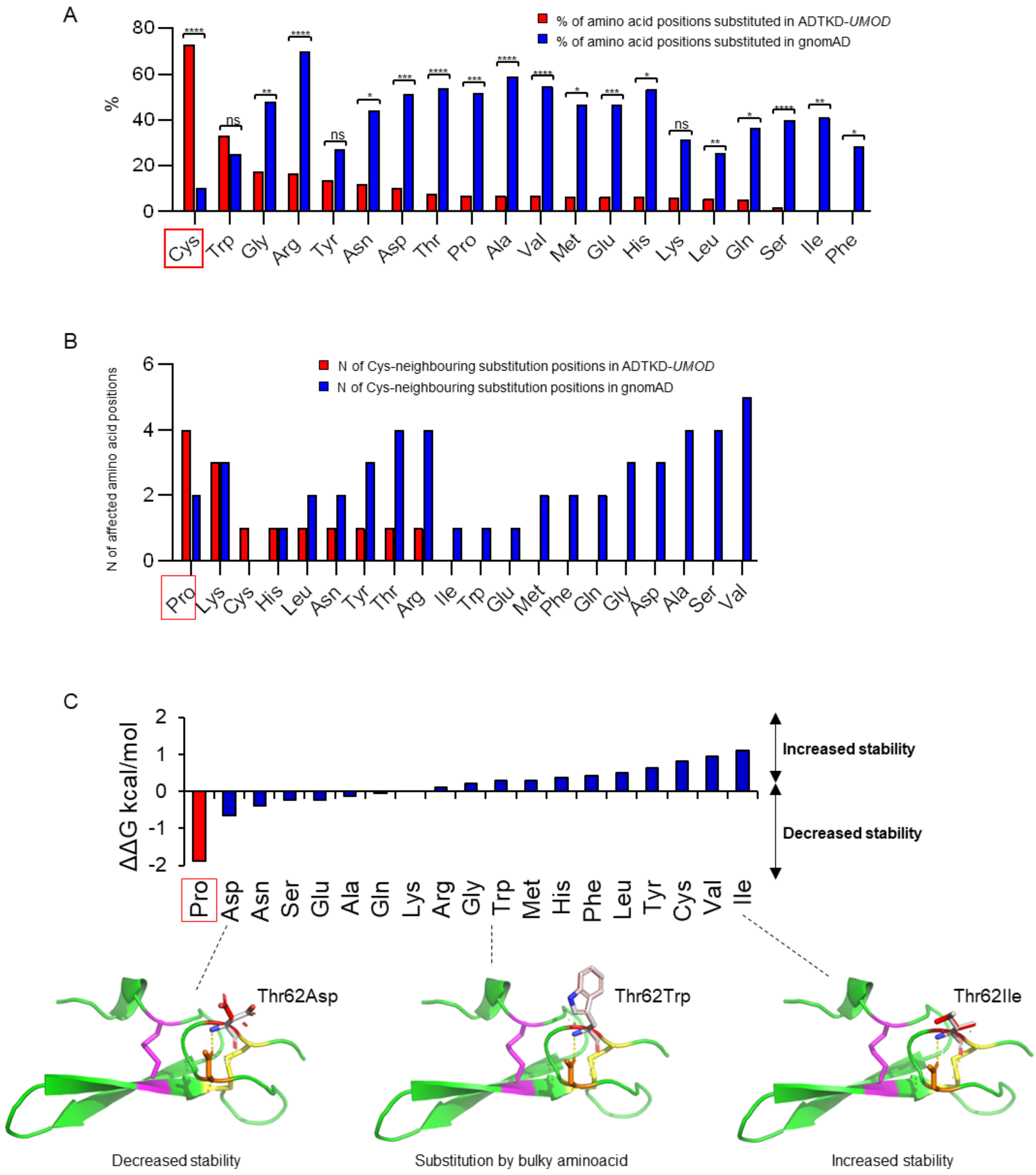

A
% of amino acid positions substituted in ADTKD-UMOD
% of amino acid positions substituted in gnomAD
%
Cys Trp Gly Arg Tyr Asn Asp Thr Pro Ala Val Met Glu His Lys Leu Gln Ser Ile Phe
B
N of Cys-neighbouring substitution positions in ADTKD-UMOD
N of Cys-neighbouring substitution positions in gnomAD
N of affected amino acid positions
Pro Lys Cys His Leu Asn Tyr Thr Arg Ile Trp Glu Met Phe Gln Gly Asp Ala Ser Val
C
ΔΔG kcal/mol
Increased stability
Decreased stability
Pro Asp Asn Ser Glu Ala Gln Lys Arg Gly Trp Met His Phe Leu Tyr Cys Val Ile
Thr62Asp
Thr62Trp
Thr62Ile
Decreased stability
Substitution by bulky aminoacid
Increased stability

**Fig. S4. UMOD amino acid substitutions and *in silico* modelling of p.Thr62 isoforms.** (A) Percentage of amino acid positions that are substituted in UMOD missense variants reported in gnomAD (blue) vs. in ADTKD-*UMOD* patients from the International ADTKD Cohort and HGMD® (red). Of note, 35/48 (73%) of cysteine positions have been substituted in patients with ADTKD-*UMOD* vs. only 5/48 (10%) positions in gnomAD. Statistical analysis using Fisher's exact test on number of substitutions in relation to total number of available positions for each amino acid; ns not significant; *p<0.05, **p<0.01, ***p<0.001, ****p<0.0001. (B) Number of amino acid substitution positions directly adjacent to cysteines in the gnomAD dataset (blue) vs. in ADTKD-*UMOD* patients from the international ADTKD cohort and HGMD® (red). Out of the total available cysteine-adjacent positions (n=94), 4 are substituted with a proline in ADTKD-*UMOD* patients, while only 2 positions are substituted with a proline in gnomAD (including p.Thr62Pro), indicating relatively poor tolerance for proline substitutions adjacent to cysteines. (C) The graph represents the double change in Gibbs free energy (ΔΔG; measure of the change in free energy between the folded and unfolded states in wild type protein and after insertion of the point mutation) as a read out of how the replacement of Thr62 with the indicated residue affects protein stability. Representative examples of modelling of UMOD EGF-like domain 1 variants at position 62. Here shown are substitutions of Thr62 with aspartic acid (as the most destabilizing mutation after proline), tryptophan (as an example of substitution with a bulky aminoacid) and isoleucine (as the most stabilizing substitution). Of note, tryptophan is the less frequent residue present in vertebrate EGF-like domains at the position corresponding to UMOD Thr62 (see Figure 3A). None of these substitutions has a strong impact on protein structure, except for p.Thr62Pro (see Figure 3B). Polar contacts are indicated by yellow dashed lines. Small green and red disks indicate the presence of atoms almost in contact and van der Waals overlap, respectively. The figure was made with PyMOL (Schrödinger LLC).

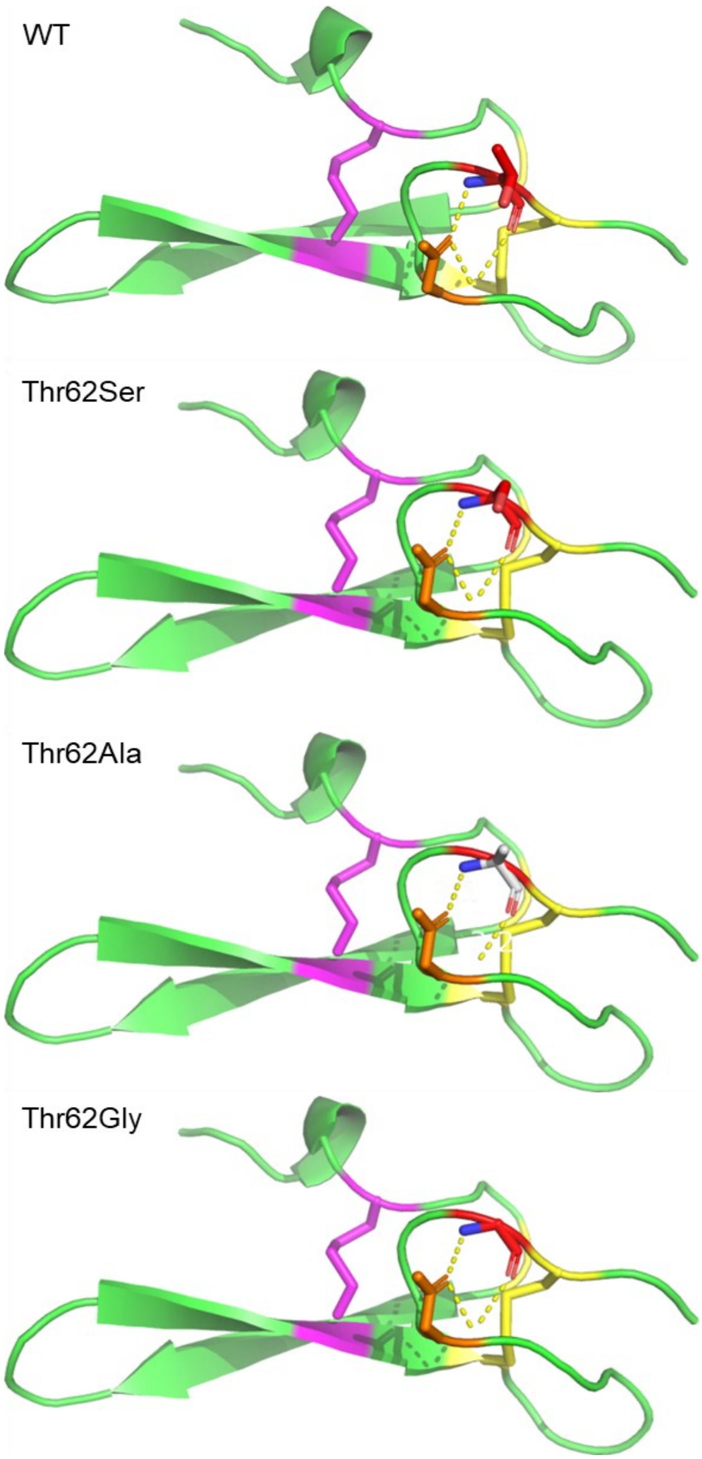


**Fig. S5. *In silico* modelling of pThr62Ser, p.Thr62Ala and p.Thr62Gly.** Modelling of the indicated UMOD isoforms. Shown here are substitutions with serine, a nucleophilic amino acid similar to proline, and two small amino acids, alanine and glycine. Polar contacts are indicated by yellow dashed line. Potential clashes in the structure are represented by red dots (not seen in any of these predictions). The figure was made with PyMOL (Schrödinger LLC).

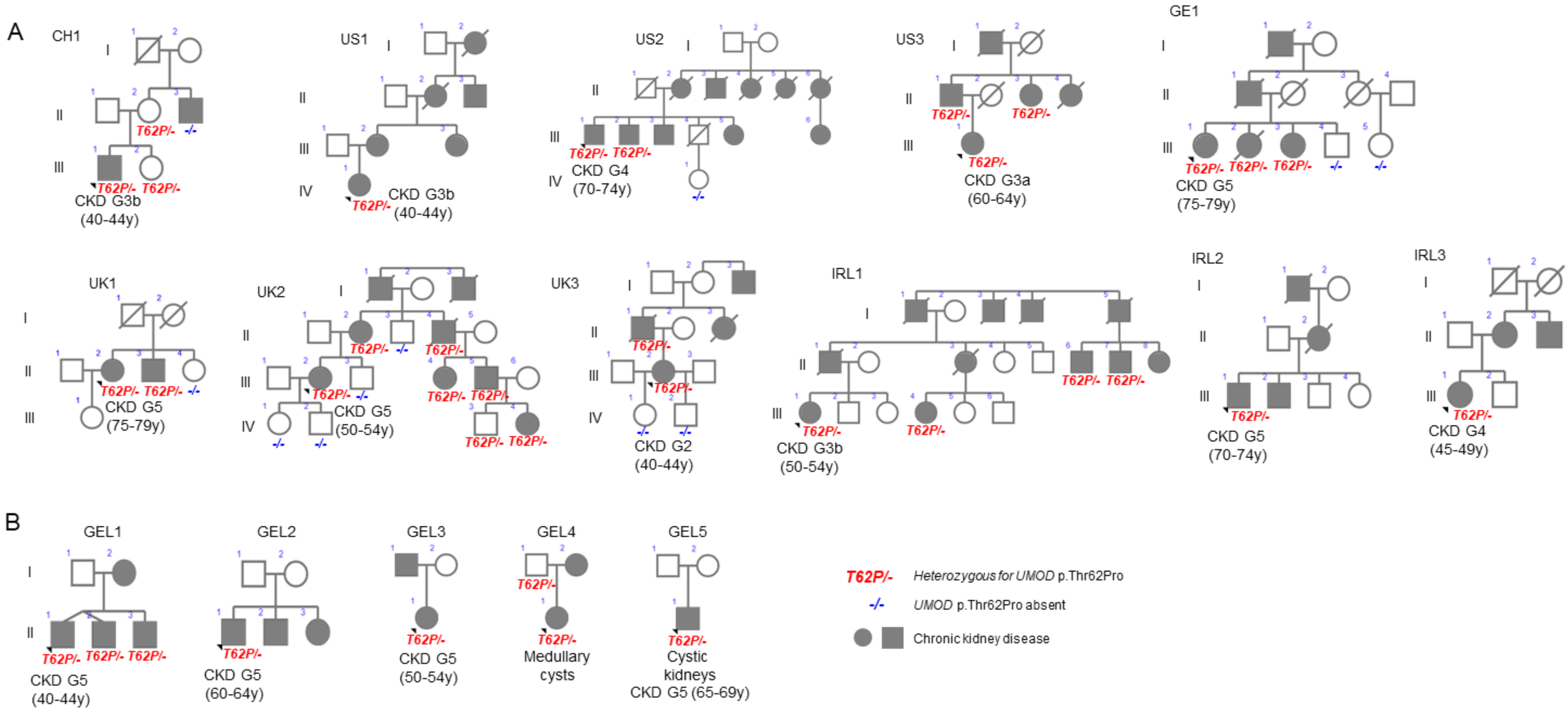


**Fig. S6. *UMOD* p.Thr62Pro genetic load in familial CKD clusters.** (A) *UMOD* p.Thr62Pro carriers in families with unexplained CKD and features compatible with autosomal dominant tubulointerstitial kidney disease identified in tertiary clinical centres and (B) identified through Genomics England 100,000 Genomes project (GEL). Patients with chronic kidney disease (CKD) are marked in grey, unaffected family members in white. The index patient is marked with an arrowhead and is labelled with CKD stadium or kidney phenotype at last evaluation and age range. For individuals with available genotype, the heterozygote status of p.Thr62Pro is marked in red, absence of *UMOD* p.Thr62Pro in blue. For full phenotype and genotype information see Table S4.

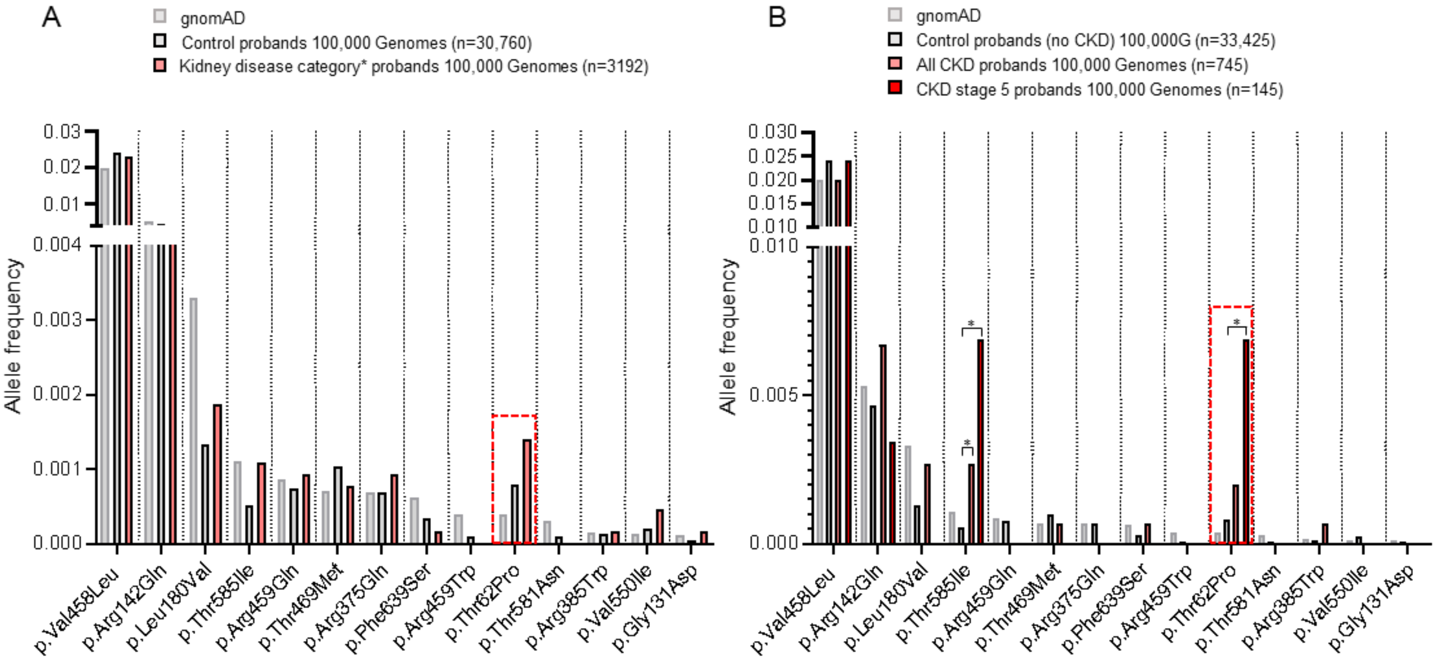


**Fig. S7. Genetic load of rare and low-frequency *UMOD* missense variants in 100,000 Genomes Project kidney disease probands.** Distribution of allelic frequencies for rare (gnomAD $10^{-3}$>AF>$10^{-4}$) or low frequency (gnomAD AF>$10^{-3}$) *UMOD* missense variants in the general population (gnomAD) and in probands enrolled in Genomics England 100,000 Genomes project. (A) 100,000 Genomes probands enrolled under any of following kidney disease categories (*): 'congenital anomalies of the kidney and the urinary tract', 'cystic kidney disease', 'familial haematuria', 'proteinuric renal disease', 'renal tract calcification', 'unexplained kidney failure in young' versus probands enrolled under any of the remaining disease categories. (B) 100,000 Genomes probands enrolled under Human Phenotype Ontology (HPO) terms 'chronic kidney disease' or 'chronic kidney disease stage 5' versus probands without chronic kidney disease. Significant enrichment using Fisher's exact test is indicated with *P<0.05.

A

| 100,000 Genomes | Allele counts (L180V/WT alleles) | AF (L180V/all alleles) | OR (95% CI) | P-value |
|---|---|---|---|---|
| **Kidney disease categories*** | | | | |
| Control probands | 82/61438 | 0.0013 | | |
| Case probands | 12/6372 | 0.0019 | 1.41 (0.76-2.52) | 0.29 |
| **HPO Chronic kidney disease** | | | | |
| Control probands | 87/66763 | 0.0013 | | |
| All CKD cases | 4/1486 | 0.0027 | 2.07 (0.80-5.21) | 0.14 |
| CKD stage 5 cases | 0/290 | 0.00 | - | - |

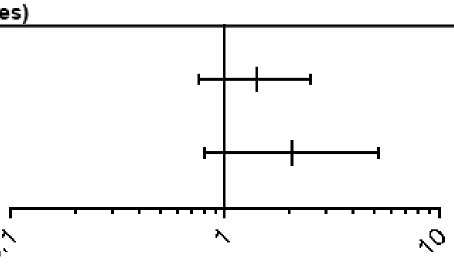


B

| 100,000 Genomes | Allele counts (T469M/WT alleles) | AF (T469M/all alleles) | OR (95% CI) | P-value |
|---|---|---|---|---|
| **Kidney disease categories*** | | | | |
| Control probands | 64/61456 | 0.00104 | | |
| Case probands | 5/6379 | 0.00078 | 0.75 (0.33-1.81) | 0.68 |
| **HPO Chronic kidney disease** | | | | |
| Control probands | 70/66780 | 0.00104 | | |
| All CKD cases | 1/1489 | 0.00067 | 0.64 (0.063-3.65) | 0.99 |
| CKD stage 5 cases | 0/290 | 0.00 | - | - |

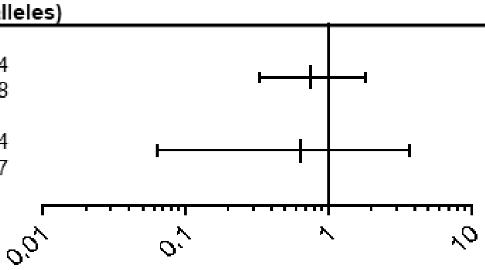


**Fig. S8. *UMOD* p.Leu180Val and p.Thr469Met in 100,000 Genomes Project kidney diseases probands and controls.** (A&B) Prevalence of p.Leu180Val and p.Thr469Met heterozygous alleles in probands from the Genomics England 100,000 Genomes project with indicated renal phenotypes and in control probands where the indicated phenotype is absent. *Kidney disease categories comprise: ‘congenital anomalies of the kidney and the urinary tract’, ‘cystic kidney disease’, ‘familial haematuria’, ‘proteinuric renal disease’, ‘renal tract calcification’, ‘unexplained kidney failure in young’. P value computed using Fisher’s exact test.

A

| | Allele counts (T62P/WT alleles) | AF (T62P/all alleles) | OR (95% CI) | P-value |
|---|---|---|---|---|
| **N183 CKD stage 3** | | | | |
| Controls | 243/306301 | 0.00079 | | |
| Cases | 2/2670 | 0.00075 | 0.94 (0.24-3.80) | >0.99 |
| **N184 CKD stage 4** | | | | |
| Controls | 245/308581 | 0.00079 | | |
| Cases | 0/390 | 0.00000 | - | - |
| **N185 CKD stage 5** | | | | |
| Controls | 143/308623 | 0.00046 | | |
| Cases | 2/328 | 0.00606 | 12.40 (3.06-50.27) | 0.012 |
| **Transplantation of kidney** | | | | |
| Controls | 243/308763 | 0.00079 | | |
| Cases | 2/208 | 0.00952 | 12.22 (3.02-49.46) | 0.012 |

B

| | Allele counts (L180V/WT alleles) | AF (L180V/all alleles) | OR (95% CI) | P-value |
|---|---|---|---|---|
| **N183 CKD stage 3** | | | | |
| Controls | 1/306543 | 3.26E-06 | | |
| Cases | 0/2672 | 0.00000 | - | - |
| **N184 CKD stage 4** | | | | |
| Controls | 1/308825 | 3.24E-06 | | |
| Cases | 0/390 | 0.00000 | - | - |
| **N185 CKD stage 5** | | | | |
| Controls | 1/308865 | 3.24E-06 | | |
| Cases | 0/350 | 0.00000 | - | - |
| **Transplantation of kidney** | | | | |
| Controls | 1/309005 | 3.24E-06 | | |
| Cases | 0/205 | 0.00000 | - | - |

C

| | Allele counts (T469M/WT alleles) | AF (T469M/all alleles) | OR (95% CI) | P-value |
|---|---|---|---|---|
| **N183 CKD stage 3** | | | | |
| Controls | 446/306090 | 0.00145 | | |
| Cases | 4/2666 | 0.00150 | 1.03 (0.39-2.76) | 0.8 |
| **N184 CKD stage 4** | | | | |
| Controls | 450/308366 | 0.00146 | | |
| Cases | 0/390 | 0.00000 | - | - |
| **N185 CKD stage 5** | | | | |
| Controls | 450/308406 | 0.00146 | | |
| Cases | 0/350 | 0.00000 | - | - |
| **Transplantation of kidney** | | | | |
| Controls | 450/308546 | 0.00146 | | |
| Cases | 0/205 | 0.00000 | - | - |

**Fig. S9. p.Thr62Pro, p.Leu180Val and p.Thr469Met and kidney disease in UK Biobank exome data.** Prevalence of p.Thr62Pro, p.Leu180Val, p.Thr469Met alleles in controls and in individuals with indicated kidney phenotypes in the UK biobank exome data (unrelated individuals with “Caucasian” genetic ancestry). P value computed using Fisher’s exact test.

**Table S1. Top 25 most common *UMOD* missense variants in gnomAD.**

| Chr | Position | rsID | REF | ALT | AA change | Allele Count | Allele Number | AF |
|---|---|---|---|---|---|---|---|---|
| 16 | 20352618 | rs55772253 | C | A | p.(Val458Leu) | 5653 | 282538 | 0.02001 |
| 16 | 20360198 | rs199835347 | C | T | p.(Arg142Gln) | 981 | 184668 | 0.00531 |
| 16 | 20360085 | rs187555378 | G | C | p.(Leu180Val) | 578 | 174180 | 0.00332 |
| 16 | 20348036 | rs111992415 | G | A | p.(Thr585Ile) | 323 | 282822 | 0.00114 |
| 16 | 20352614 | rs201761378 | C | T | p.(Arg459Gln) | 247 | 282566 | 0.00087 |
| 16 | 20352584 | rs143583842 | G | A | p.(Thr469Met) | 203 | 282716 | 0.00072 |
| 16 | 20357506 | rs151061376 | C | T | p.(Arg375Gln) | 196 | 282574 | 0.00069 |
| 16 | 20344643 | rs145165861 | A | G | p.(Phe639Ser) | 177 | 281046 | 0.00063 |
| 16 | 20352615 | rs139607138 | G | A | p.(Arg459Trp) | 109 | 282528 | 0.00039 |
| 16 | 20360439 | rs143248111 | T | G | p.(Thr62Pro) | 99 | 282046 | 0.00035 |
| 16 | 20348048 | rs143641292 | G | T | p.(Thr581Asn) | 84 | 282786 | 0.00030 |
| 16 | 20357477 | rs200414962 | G | A | p.(Arg385Trp) | 41 | 282438 | 0.00015 |
| 16 | 20348705 | rs188709583 | C | T | p.(Val550Ile) | 36 | 282824 | 0.00013 |
| 16 | 20360231 | rs368943553 | C | T | p.(Gly131Asp) | 23 | 202250 | 0.00011 |
| 16 | 20360193 | rs769398465 | C | T | p.(Asp144Asn) | 17 | 183688 | 0.00009 |
| 16 | 20357449 | rs532447307 | G | A | p.(Thr394Met) | 25 | 282444 | 0.00009 |
| 16 | 20352618 | rs55772253 | C | G | p.(Val458Leu) | 25 | 282538 | 0.00009 |
| 16 | 20359938 | rs756226236 | T | A | p.(Met229Leu) | 19 | 229628 | 0.00008 |
| 16 | 20348027 | rs387907549 | C | T | p.(Arg588Gln) | 22 | 282850 | 0.00008 |
| 16 | 20357476 | rs141355380 | C | T | p.(Arg385Gln) | 19 | 251092 | 0.00008 |
| 16 | 20359889 | rs753492646 | C | T | p.(Arg245His) | 16 | 216168 | 0.00007 |
| 16 | 20359781 | rs543775691 | T | C | p.(Glu281Gly) | 19 | 278806 | 0.00007 |
| 16 | 20352446 | rs550521976 | G | A | p.(Thr515Met) | 19 | 282720 | 0.00007 |
| 16 | 20352542 | rs779351986 | G | A | p.(Ala483Val) | 15 | 251422 | 0.00006 |
| 16 | 20359865 | rs760253448 | C | T | p.(Gly253Asp) | 15 | 258220 | 0.00006 |

Variants denoting non-canonical transcripts have been removed and consequences have been checked for consistency with *UMOD* transcript ENST00000302509.8.

**Table S2. Top predicted pathogenic *UMOD* missense variants in gnomAD (REVEL score >0.75).**

| Location | Allele | HGVSc | HGVSp | SIFT | Condel | PolyPhen | REVEL | ClinVar | gnomAD AC |
|---|---|---|---|---|---|---|---|---|---|
| 16:20359916 | A | c.707C>T | p.Pro236Leu | deleterious(0) | deleterious(0.945) | probably_damaging(1) | 0.973 | P/LP | 1 |
| 16:20359802 | C | c.821A>G | p.Tyr274Cys | deleterious(0) | deleterious(0.935) | probably_damaging(0.999) | 0.955 | - | 1 |
| 16:20360264 | T | c.359G>A | p.Cys120Tyr | deleterious(0) | deleterious(0.935) | probably_damaging(0.999) | 0.954 | - | 1 |
| 16:20348774 | G | c.1579T>C | p.Cys527Arg | deleterious(0) | deleterious(0.945) | probably_damaging(1) | 0.930 | - | 1 |
| 16:20359829 | A | c.794A>T | p.Lys265Met | deleterious(0) | deleterious(0.919) | probably_damaging(0.998) | 0.909 | - | 1 |
| 16:20360306 | A | c.317G>T | p.Cys106Phe | deleterious(0) | deleterious(0.935) | probably_damaging(0.999) | 0.906 | Conflicting | 2 |
| 16:20352482 | C | c.1508T>G | p.Met503Arg | deleterious(0) | deleterious(0.867) | probably_damaging(0.979) | 0.886 | - | 1 |
| 16:20357461 | T | c.1169G>A | p.Gly390Glu | deleterious(0) | deleterious(0.919) | probably_damaging(0.998) | 0.882 | - | 1 |
| 16:20348656 | T | c.1697G>A | p.Cys566Tyr | deleterious(0) | deleterious(0.945) | probably_damaging(1) | 0.874 | - | 1 |
| 16:20348659 | C | c.1694A>G | p.His565Arg | deleterious(0) | deleterious(0.945) | probably_damaging(1) | 0.862 | - | 1 |
| 16:20359902 | T | c.721G>A | p.Gly241Ser | deleterious(0.02) | deleterious(0.883) | probably_damaging(1) | 0.860 | - | 1 |
| 16:20357644 | G | c.986T>C | p.Leu329Pro | deleterious(0) | deleterious(0.911) | probably_damaging(0.997) | 0.843 | - | 1 |
| 16:20357626 | T | c.1004G>A | p.Cys335Tyr | deleterious(0) | deleterious(0.945) | probably_damaging(1) | 0.834 | - | 1 |
| 16:20360507 | A | c.116C>T | p.Ala39Val | deleterious(0) | deleterious(0.919) | probably_damaging(0.998) | 0.829 | - | 1 |
| 16:20360231 | T | c.392G>A | p.Gly131Asp | deleterious(0) | deleterious(0.902) | probably_damaging(0.995) | 0.822 | B | 23 |
| 16:20359943 | A | c.680C>T | p.Ala227Val | deleterious(0.01) | deleterious(0.835) | probably_damaging(0.983) | 0.819 | - | 1 |
| 16:20348662 | T | c.1691T>A | p.Leu564Gln | deleterious(0) | deleterious(0.945) | probably_damaging(1) | 0.813 | - | 1 |
| 16:20348684 | T | c.1669G>A | p.Gly557Arg | deleterious(0.01) | deleterious(0.905) | probably_damaging(1) | 0.802 | - | 1 |
| 16:20359873 | T | c.750C>A | p.His250Gln | deleterious(0.02) | deleterious(0.561) | possibly_damaging(0.527) | 0.801 | - | 1 |
| 16:20359826 | A | c.797C>T | p.Ala266Val | deleterious(0) | deleterious(0.889) | probably_damaging(0.991) | 0.798 | VUS | 3 |
| 16:20348759 | A | c.1594G>T | p.Asp532Tyr | deleterious(0) | deleterious(0.945) | probably_damaging(1) | 0.794 | - | 1 |
| 16:20357552 | T | c.1078T>A | p.Tyr360Asn | tolerated(0.05) | deleterious(0.820) | probably_damaging(0.998) | 0.794 | - | 1 |
| 16:20348638 | C | c.1715A>G | p.Asp572Gly | deleterious(0) | deleterious(0.902) | probably_damaging(0.995) | 0.788 | - | 2 |
| 16:20352569 | A | c.1421G>T | p.Gly474Val | deleterious(0) | deleterious(0.935) | probably_damaging(0.999) | 0.788 | - | 1 |

| | | | | | | | | | |
|---|---|---|---|---|---|---|---|---|---|
| 16:20359818 | T | c.805G>A | p.Gly269Ser | deleterious(0.01) | deleterious(0.626) | possibly_damaging(0.63) | 0.788 | - | 1 |
| 16:20352630 | T | c.1360G>A | p.Gly454Ser | deleterious(0) | deleterious(0.945) | probably_damaging(1) | 0.779 | - | 3 |
| 16:20352636 | T | c.1354G>A | p.Gly452Arg | deleterious(0) | deleterious(0.935) | probably_damaging(0.999) | 0.779 | - | 8 |
| 16:20352554 | G | c.1436T>C | p.Leu479Pro | deleterious(0) | deleterious(0.945) | probably_damaging(1) | 0.760 | - | 1 |
| 16:20360297 | T | c.326T>A | p.Val109Glu | deleterious(0) | deleterious(0.871) | probably_damaging(0.982) | 0.759 | LP | 3 |
| 16:20357554 | C | c.1076T>G | p.Met359Arg | tolerated(0.14) | deleterious(0.484) | possibly_damaging(0.664) | 0.758 | - | 2 |

UMOD transcript ENST00000302509.4. Variants detected in ADTKD patients are shaded in grey. SIFT, Sorting Intolerant From Tolerant; Condel, CONsensus DELeteriousness; REVEL, rare exome variant ensemble learner (27) (a score >0.75 corresponds to a sensitivity of ~0.5 and a specificity of ~0.95 for pathogenic variants in the training dataset). ClinVar last accessed 06.2021, B, benign; P, pathogenic; LP, likely pathogenic; VUS, variant of unknown significance. AC, allele count.

**Table S3. Missense *UMOD* variants reported in ADTKD families.**

| Nr | Exon | Domain | Nucleotide Change | Effect on Coding Sequence | Families ADTKD Cohort | Phenotype HGMD® | Ref paper HGMD® | families HGMD® | Families total |
|---|---|---|---|---|---|---|---|---|---|
| 1 | 3 | I | c.95G>A | p.(Cys32Tyr) | 1 | FJHN/MCKD syndrome | (Vylet'al et al., 2006) (28) | 1 | **2** |
| 2 | 3 | I | c.96C>G | p.(Cys32Trp) | 1 | Hyperuricaemic nephropathy, juvenile | (Williams et al., 2009) (29) | 1 | **2** |
| 3 | 3 | I | c.100G>A | p.(Glu34Lys) | 1 | Tubulointerstitial nephritis | (Bollée et al., 2011) (12) | 1 | **1*** |
| 4 | 3 | I | c.115G>A | p.(Ala39Thr) | 1 | | | 0 | **1** |
| 5 | 3 | I | C.122G>A | p.(Cys41Tyr) | 1 | | | 0 | **1** |
| 6 | 3 | I | c.149G>C | p.(Cys50Ser) | 0 | Reduced kidney function, hyperuricaemia and impaired urine-concentrating ability | (Benetti et al., 2009) (30) | 1 | **1** |
| 7 | 3 | I | c.155G>A | p.(Cys52Tyr) | 1 | Chronic kidney disease, autosomal dominant | (Ekici et al., 2014) (7) | 1 | **2** |
| 8 | 3 | I | c.155G>C | p.(Cys52Ser) | 1 | | | 0 | **1** |
| 9 | 3 | I | c.156T>G | p.(Cys52Trp) | 0 | Hyperuricaemic nephropathy, juvenile | (Kudo et al., 2004) (31) | 1 | **1** |
| 10 | 3 | I | c.163G>A | p.(Gly55Ser) | 1 | | | 0 | **1** |
| 11 | 3 | I | c.172G>T | p.(Gly58Cys) | 0 | Uromodulin-associated kidney disease | (Zaucke et al., 2010) (32) | 1 | **1** |

| | | | | | | | | | |
|---|---|---|---|---|---|---|---|---|---|
| 12 | 3 | I | c.176A>C | p.(Asp59Ala) | 1 | Hyperuricaemic nephropathy, juvenile | (Dahan et al., 2003) (5) | 1 | **1*** |
| 13 | 3 | I | c.184A>C | p.(Thr62Pro) | 1 | Chronic kidney disease, autosomal dominant? | (Ekici et al., 2014) (7) | 1 | **2** |
| 14 | 3 | I | c.188G>C | p.(Cys63Ser) | 1 | | | 0 | **1** |
| 15 | 3 | I | c.187T>C | p.(Cys63Arg) | 0 | Hyperuricaemic nephropathy, juvenile | (M.-N. Lee et al., 2013) (33) | 1 | **1** |
| 16 | 3 | I | c.189C>G | p.(Cys63Trp) | 0 | Hyperuricaemic nephropathy, juvenile | (Alaygut et al., 2013) (34) | 1 | **1** |
| 17 | 3 | II | c.202G>A | p.(Glu68Lys) | 1 | | | 0 | **1** |
| 18 | 3 | II | c.206G>A | p.(Cys69Tyr) | 0 | Uromodulin-associated kidney disease | (Zaucke et al., 2010) (32) | 1 | **1** |
| 19 | 3 | II | c.205T>C | p.(Cys69Arg) | 1 | Tubulointerstitial nephritis | (Bollée et al., 2011) (12) | 1 | **1*** |
| 20 | 3 | II | c.228C>G; c.229T>G | p.(Asn76Lys) p.(Cys77Gly) | 1 | | | 0 | **1** |
| 21 | 3 | II | c.229T>G | p.(Cys77Gly) | 0 | FJHN/MCKD syndrome | (Wolf et al., 2007) (35) | 1 | **1** |
| 22 | 3 | II | c.229T>C | p.(Cys77Arg) | 1 | | | 0 | **1** |
| 23 | 3 | II | c.230G>A | p.(Cys77Tyr) | 2 | Hyperuricaemic nephropathy, juvenile | (Turner et al., 2003) (36) | 1 | **3** |
| 24 | 3 | II | c.247T>C | p.(Cys83Arg) | 1 | | | 0 | **1** |
| 25 | 3 | II | c.254A>G | p.(Asn85Ser) | 1 | | | 0 | **1** |
| 26 | 3 | II | c.255C>G | p.(Asn85Lys) | 2 | | | 0 | **2** |

| | | | | | | | | | |
|---|---|---|---|---|---|---|---|---|---|
| 27 | 3 | II | c.274T>C | p.(Cys92Arg) | 1 | | | 0 | **1** |
| 28 | 3 | II | c.274T>G | p.(Cys92Gly) | 1 | | | 0 | **1** |
| 29 | 3 | II | c.275G>A | p.(Cys92Tyr) | 2 | | | 0 | **2** |
| 30 | 3 | II | c.282C>G | p.(Cys94Trp) | 1 | | | 0 | **1** |
| 31 | 3 | II | c.307G>T | p.(Gly103Cys) | 1 | Medullary cystic kidney disease 2 | (Hart et al., 2002) (37) | 1 | **1*** |
| 32 | 3 | II | c.317G>T | p.(Cys106Phe) | 9 | | | 0 | **9** |
| 33 | 3 | II | c.316T>G | p.(Cys106Gly) | 2 | | | 0 | **2** |
| 34 | 3 | II | c.317G>A | p.(Cys106Tyr) | 1 | Uromodulin-associated kidney disease | (Zaucke et al., 2010) (32) | 1 | **2** |
| 35 | 3 | III | c.326T>A | p.(Val109Glu) | 0 | Hyperuricaemic nephropathy, juvenile | (Liu et al., 2013) (38) | 1 | **1** |
| 36 | 3 | III | c.334T>C | p.(Cys112Arg) | 1 | Hyperuricaemic nephropathy, juvenile | (Dahan et al., 2003) (5) | 1 | **1*** |
| 37 | 3 | III | c.358T>C | p.(Cys120Arg) | 1 | | | 0 | **1** |
| 38 | 3 | III | c.376T>C | p.(Cys126Arg) | 1 | Hyperuricaemic nephropathy, juvenile | (Turner et al., 2003) (36) | 1 | **2** |
| 39 | 3 | III | c.383A>G | p.(Asn128Ser) | 4 | Hyperuricaemic nephropathy, juvenile | (Turner et al., 2003) (36) | 1 | **5** |
| 40 | 3 | III | c.404G>A | p.(Cys135Tyr) | 2 | | | 0 | **2** |
| 41 | 3 | III | c.403T>A | p.(Cys135Ser) | 1 | Hyperuricaemic nephropathy, juvenile | (Kudo et al., 2004) (31) | 1 | **2** |
| 42 | 3 | III | c.404G>T | p.(Cys135Phe) | 2 | | | 0 | **2** |
| 43 | 3 | III | c.405C>G | p.(Cys135Trp) | 1 | | | 0 | **1** |

| | | | | | | | | | |
|---|---|---|---|---|---|---|---|---|---|
| 44 | 3 | III | c.403T>G | p.(Cys135Gly) | 0 | Uromodulin-associated kidney disease | (Kuma et al., 2015) (39) | 1 | **1** |
| 45 | 3 | III | c.410G>A | p.(Cys137Tyr) | 0 | Hyperuricaemic nephropathy, juvenile | (Taylor et al., 2015) (40) | 1 | **1** |
| 46 | 3 | III | c.442T>C | p.(Cys148Arg) | 2 | Tubulointerstitial nephritis | (Bollée et al., 2011) (12) | 1 | **2*** |
| 47 | 3 | III | c.442T>A, c.443G>C | p.(Cys148Ser) | 2 | | | 0 | **2** |
| 48 | 3 | III | c.443G>A | p.(Cys148Tyr) | 4 | Hyperuricaemic nephropathy, juvenile | (Hart et al., 2002) (37) | 1 | **4*** |
| 49 | 3 | III | c.444T>G | p.(Cys148Trp) | 0 | Medullary cystic kidney disease 2 | (Rampoldi et al., 2003) (41) | 1 | **1** |
| 50 | 3 | | c.449G>C | p.(Cys150Ser) | 1 | Medullary cystic kidney disease 2 | (Rampoldi et al., 2003) (41) | 1 | **2** |
| 51 | 3 | | c.448T>A | p.(Cys150Ser) | 0 | Uromodulin-associated kidney disease | (Zaucke et al., 2010) (32) | 1 | **1** |
| 52 | 3 | | c.463T>C | p.(Cys155Arg) | 0 | Chronic kidney disease | (Stewart et al., 2015) (42) | 1 | **1** |
| 53 | 3 | | c.478G>C | p.(Asp160His) | 1 | | | 0 | **1** |
| 54 | 3 | | c.509G>A | p.(Cys170Tyr) | 2 | Hyperuricaemic nephropathy, juvenile | (Dahan et al., 2003) (5) | 1 | **2*** |
| 55 | 3 | | c.514G>C | p.(Asp172His) | 2 | Tubulointerstitial nephritis | (Bollée et al., 2011) (12) | 1 | **2*** |
| 56 | 3 | | c.518C>T | p.(Pro173Leu) | 1 | | | 0 | **1** |

| | | | | | | | | | |
|---|---|---|---|---|---|---|---|---|---|
| 57 | 3 | | c.518C>G | p.(Pro173Arg) | 0 | Hyperuricaemic nephropathy, juvenile | (Iguchi et al., 2013) (43) | 1 | **1** |
| 58 | 3 | | c.520T>C | p.(Cys174Arg) | 2 | Uromodulin-associated kidney disease | (Moskowitz et al., 2013) (44) | 1 | **3** |
| 59 | 3 | | c.533G>C | p.(Arg178Pro) | 14 | | | 0 | **14** |
| 60 | 3 | | c.539T>C | p.(Leu180Pro) | 2 | | | 0 | **2** |
| 61 | 3 | | c.538C>G | p.(Leu180Val) | 1 | | | 0 | **1** |
| 62 | 3 | | c.552G>C | p.(Trp184Cys) | 1 | Tubulointerstitial nephritis | (Bollée et al., 2011) (12) | 1 | **1*** |
| 63 | 3 | | c.553C>A | p.(Arg185Ser) | 1 | Hyperuricaemic nephropathy, juvenile | (Dahan et al., 2003) (5) | 1 | **1*** |
| 64 | 3 | | c.554G>A | p.(Arg185His) | 2 | Tubulointerstitial nephritis | (Bollée et al., 2011) (12) | 1 | **2*** |
| 65 | 3 | | c.553C>T | p.(Arg185Cys) | 4 | Tubulointerstitial nephritis | (Bollée et al., 2011) (12) | 1 | **4*** |
| 66 | 3 | | c.553C>G | p.(Arg185Gly) | 1 | Hyperuricaemic nephropathy, juvenile | (Bleyer et al., 2004) (45) | 1 | **1*** |
| 67 | 3 | | c.584G>A | p.(Cys195Tyr) | 1 | Hyperuricaemic nephropathy, juvenile | (Spain et al., 2014) (46) | 1 | **2** |
| 68 | 3 | | c.584G>T | p.(Cys195Phe) | 0 | Hyperuricaemic nephropathy, juvenile | (Kudo et al., 2004) (31) | 1 | **1** |
| 69 | 3 | | c.585_586CG>TA | p.(Asp196Asn) | 1 | | | 0 | **1** |
| 70 | 3 | | c.586G>T | p.(Asp196Tyr) | 2 | Hyperuricaemic nephropathy, juvenile | (Lhotta et al., 2009) (47) | 1 | **3** |
| 71 | 3 | | c.586G>A | p.(Asp196Asn) | 0 | Hyperuricaemic nephropathy, juvenile | (Williams et al., 2009) (29) | 1 | **1** |

| | | | | | | | | | |
|---|---|---|---|---|---|---|---|---|---|
| 72 | 3 | D8C | c.602G>A | p.(Gly201Asp) | 1 | | | 0 | **1** |
| 73 | 3 | D8C | c.605G>C | p.(Trp202Ser) | 1 | Hyperuricaemic nephropathy, juvenile | (Kudo et al., 2004) (31) | 1 | **2** |
| 74 | 3 | D8C | c.606G>T | p.(Trp202Cys) | 1 | | | 0 | **1** |
| 75 | 3 | D8C | c.607T>G | p.(Tyr203Asp) | 1 | | | 0 | **1** |
| 76 | 3 | D8C | c.611G>C | p.(Arg204Pro) | 1 | | | 0 | **1** |
| 77 | 3 | D8C | c.610C>G | p.(Arg204Gly) | 4 | Hyperuricaemic nephropathy, juvenile | (Dahan et al., 2003) (5) | 1 | **4*** |
| 78 | 3 | D8C | c.628G>A | p.(Gly210Ser) | 2 | Tubulointerstitial nephritis | (Bollée et al., 2011) (12) | 2 | **2*** |
| 79 | 3 | D8C | c.634C>T | p.(Arg212Cys) | 0 | Uromodulin-associated kidney disease | (Schaeffer et al., 2012) (48) | 1 | **1** |
| 80 | 3 | D8C | c.649T>G | p.(Cys217Gly) | 1 | Hyperuricaemic nephropathy, juvenile | (Dahan et al., 2003) (5) | 1 | **1*** |
| 81 | 3 | D8C | c.651C>G | p.(Cys217Trp) | 0 | Hyperuricaemic nephropathy, juvenile | (Williams et al., 2009) (29) | 1 | **1** |
| 82 | 3 | D8C | c.649T>C | p.(Cys217Arg) | 1 | Hyperuricaemic nephropathy, juvenile | (Hart et al., 2002) (37) | 1 | **1*** |
| 83 | 3 | D8C | c.665G>C | p.(Arg222Pro) | 1 | Hyperuricaemic nephropathy, juvenile | (Dahan et al., 2003) (5) | 1 | **1*** |
| 84 | 3 | D8C | c.668G>A | p.(Cys223Tyr) | 2 | Hyperuricaemic nephropathy, juvenile | (Bleyer et al., 2003) (49) | 1 | **2*** |
| 85 | 3 | D8C | c.667T>C | p.(Cys223Arg) | 0 | Hyperuricaemic nephropathy, juvenile | (Williams et al., 2009) | 1 | **1** |

| | | | | | | | | | |
|---|---|---|---|---|---|---|---|---|---|
| | | | | | | | (29) | | |
| 86 | 3 | D8C | c.674C>T | p.(Thr225Met) | 4 | Hyperuricaemic nephropathy, juvenile | (Dahan et al., 2003) (5) | 1 | **4*** |
| 87 | 3 | D8C | c.674C>A | p.(Thr225Lys) | 1 | Medullary cystic kidney disease 2 | (Wolf et al., 2003) (50) | 1 | **2** |
| 88 | 3 | D8C | c.686T>G | p.(Met229Arg) | 1 | FJHN/MCKD syndrome | (Vylet'al et al., 2006) (28) | 1 | **2** |
| 89 | 3 | D8C | c.688T>C | p.(Trp230Arg) | 1 | Uromodulin-associated kidney disease | (Zaucke et al., 2010) (32) | 1 | **2** |
| 90 | 3 | D8C | c.706C>T | p.(Pro236Ser) | 2 | | | 0 | **2** |
| 91 | 3 | D8C | c.707C>G | p.(Pro236Arg) | 1 | Hyperuricaemic nephropathy, juvenile | (Bernascone et al., 2006) (51) | 1 | **2** |
| 92 | 3 | D8C | c.707C>T | p.(Pro236Leu) | 6 | Hyperuricaemic nephropathy, juvenile | (Kudo et al., 2004) (31) | 1 | **7** |
| 93 | 3 | D8C | c.707C>A | p.(Pro236Gln) | 0 | Hyperuricaemic nephropathy, juvenile | (Liu et al., 2013) (38) | 1 | **1** |
| 94 | 3 | D8C | c.710C>G | p.(Ser237Cys) | 1 | Tubulointerstitial nephritis | (Bollée et al., 2011) (12) | 1 | **1*** |
| 95 | 3 | D8C | c.744C>G | p.(Cys248Trp) | 2 | Medullary cystic kidney disease 2 | (Wolf et al., 2003) (50) | 1 | **3** |
| 96 | 3 | D8C | c.743G>C | p.(Cys248Ser) | 0 | Uromodulin-associated kidney disease | (Zaucke et al., 2010) (32) | 1 | **1** |
| 97 | 3 | D8C | c.749A>T | p.(His250Leu) | 1 | Tubulointerstitial nephritis | (Bollée et al., 2011) (12) | 1 | **1*** |

| | | | | | | | | | |
|---|---|---|---|---|---|---|---|---|---|
| 98 | 3 | D8C | c.750C>A | p.(His250Gln) | 0 | Tubulointerstitial nephropathy and hyperuricaemia | (Raffler et al., 2016) (52) | 1 | **1** |
| 99 | 3 | D8C | c.757G>T | P.(Gly253Cys) | 1 | | | 0 | **1** |
| 100 | 3 | D8C | c.764G>A | p.(Cys255Tyr) | 0 | Hyperuricaemic nephropathy, juvenile | (Turner et al., 2003) (36) | 1 | **1** |
| 101 | 3 | D8C | c.767G>A | p.(Cys256Tyr) | 0 | Uromodulin-associated kidney disease | (Schaeffer et al., 2012) (48) | 1 | **1** |
| 102 | 3 | D8C | c.770T>C | p.(Leu257Pro) | 2 | | | 0 | **2** |
| 103 | 3 | D8C | c.774G>C | p.(Trp258Cys) | 1 | | | 0 | **1** |
| 104 | 3 | D8C | c.800G>T | p.(Cys267Phe) | 1 | | | 0 | **1** |
| 105 | 3 | D8C | c.805G>T | p.(Gly269Cys) | 1 | | | 0 | **1** |
| 106 | 3 | D8C | c.808G>T | p.(Gly270Cys) | 1 | Uromodulin-associated kidney disease | (Nasr et al., 2008) (53) | 1 | **2** |
| 107 | 3 | D8C | c.817G>C | p.(Val273Leu) | 1 | | | 0 | **1** |
| 108 | 3 | D8C | c.817G>T | p.(Val273Phe) | 0 | FJHN/MCKD syndrome | (Vylet'al et al., 2006) (28) | 1 | **1** |
| 109 | 3 | D8C | c.820T>C | p.(Tyr274His) | 1 | | | 0 | **1** |
| 110 | 3 | D8C | c.821A>G | p.(Tyr274Cys) | 0 | Uromodulin-associated kidney disease | (Zaucke et al., 2010) (32) | 1 | **1** |
| 111 | 3 | D8C | c.844T>C | p.(Cys282Arg) | 1 | Hyperuricaemic nephropathy, juvenile | (Dahan et al., 2003) (5) | 1 | **1*** |
| 112 | 3 | D8C | c.844T>A | p.(Cys282Ser) | 1 | | | 0 | **1** |

| | | | | | | | | | |
|---|---|---|---|---|---|---|---|---|---|
| 113 | 3 | D8C | c.851T>C | p.(Leu284Pro) | 2 | | | 0 | **2** |
| 114 | 3 | D8C | c.854C>A | p.(Ala285Glu) | 3 | | | 0 | **3** |
| 115 | 3 | D8C | c.857A>G | p.(Tyr286Cys) | 2 | | | 0 | **2** |
| 116 | 3 | D8C | c.860G>A | p.(Cys287Tyr) | 1 | | | 0 | **1** |
| 117 | 4 | IV | c.890G>A | p.(Cys297Tyr) | 1 | Tubulointerstitial nephritis | (Bollée et al., 2011) (12) | 1 | **1*** |
| 118 | 4 | IV | c.891T>G | p.(Cys297Trp) | 1 | Renal insufficiency | (Schäffer et al., 2010) (54) | 1 | **2** |
| 119 | 4 | IV | c.898T>G | p.(Cys300Gly) | 1 | Hyperuricaemic nephropathy, juvenile | (Turner et al., 2003) (36) | 1 | **2** |
| 120 | 4 | IV | c.899G>A | p.(Cys300Tyr) | 0 | Medullary cystic kidney disease 2 | (Bleyer et al., 2005) (55) | 1 | **1** |
| 121 | 4 | IV | c.898T>A | p.(Cys300Ser) | 1 | | | 0 | **1** |
| 122 | 4 | IV | c.920A>C | p.(Lys307Thr) | 0 | Hyperuricaemic nephropathy, juvenile | (Calado et al., 2005) (56) | 1 | **1** |
| 123 | 4 | IV | c.943T>C | p.(Cys315Arg) | 2 | Glomerulocystic kidney disease | (Rampoldi et al., 2003) (41) | 1 | **3** |
| 124 | 4 | IV | c.944G>A | p.(Cys315Tyr) | 2 | Tubulointerstitial nephritis | (Bollée et al., 2011) (12) | 3 | **3*** |
| 125 | 4 | IV | c.947A>C | p.(Gln316Pro) | 1 | Hyperuricaemic nephropathy, juvenile | (Lens et al., 2005) (57) | 2 | **3** |
| 126 | 4 | IV | c.950G>A | p.(Cys317Tyr) | 1 | Medullary cystic kidney disease 2 | (Rampoldi et al., 2003) (41) | 1 | **2** |

| | | | | | | | | | |
|---|---|---|---|---|---|---|---|---|---|
| 127 | 5 | ZP_N | c.1039T>G | p.(Cys347Gly) | 0 | Hyperuricaemic nephropathy, juvenile | (Tinschert et al., 2004) (58) | 1 | **1** |
| 128 | 5 | ZP_N | c.1039T>C | p.(Cys347Arg) | 0 | Congenital anomalies of the kidney and urinary tract | (Nicolaou et al., 2016) (59) | 1 | **1** |
| 129 | 7 | IHP | c.1382C>A | p.(Ala461Glu) | 1 | Hyperuricaemic nephropathy, juvenile | (D. H. Lee et al., 2010) (60) | 1 | **2** |
| 130 | 7 | ZP_C | c.1406C>T | p.(Thr469Met) | 2 | Tubulointerstitial nephritis | (Bollée et al., 2011) (12) | 1 | **2*** |
| 131 | 7 | ZP_C | c.1462G>C | p.(Gly488Arg) | 1 | Hyperuricaemic nephropathy, juvenile | (Williams et al., 2009) (29) | 1 | **2** |
| 132 | 9 | CTP | c.1813A>G | p.(Thr605Ala) | 0 | Hyperuricaemic nephropathy, juvenile | (Wei et al., 2012) (61) | 1 | 1 |

*HGMD® entries correspond to patients included in the International ADTKD Cohort
86 mutations from HGMD® database (Version 2019.1, updated in March 2019) and 101 mutations from the International ADTKD Cohort (Belgo-Swiss and Wake Forest). *UMOD* transcript: ENST00000302509. Abbreviations as follows: FJHN, familial juvenile hyperuricaemic nephropathy; MCKD, medullary cystic kidney disease.

**Table S4. Characteristics of *UMOD* p.Thr62Pro carriers in families with unexplained CKD.**

| Fam | Patient | Genotyping | Genotype | Phenotype |
|---|---|---|---|---|
| CH1 | III.1 (index) | Tubulopathy gene panel | *UMOD* p.T62P (+) | CKD G3b (40-44y), minimal proteinuria |
| | | Cystic kidney disease panel | | low FEUA with gout |
| | | MLPA *HNF1B* | | Small kidneys with cortical cysts |
| | | *MUC1* VNTR sequencing + MS | | |
| | III.2 | *UMOD* Sanger sequencing | *UMOD* p.T62P (+) | unaffected |
| | II.2 | *UMOD* Sanger sequencing | *UMOD* p.T62P (+) | Unaffected (70-74y): normal eGFR and serum electrolytes, no proteinuria, normal uric acid, normal sonography of kidneys |
| | II.3 | *UMOD* Sanger sequencing | *UMOD* p.T62P (-) | CKD G5 (45-49y), proteinuria (2.9g/d) arterial hypertension |
| US1 | IV.1 (index) | *UMOD* Sanger sequencing (exons 3,4) | *UMOD* p.T62P (+) | CKD G3b (40-44y) |
| | | *REN* Sanger sequencing | | |
| | | *MUC1* VNTR sequencing + MS | | |
| | III.2 | / | / | CKD G5 (50-54y) |
| | III.3 | / | / | CKD |
| | II.2 | / | / | CKD G5 (early 50s) |
| | II.3 | / | / | kidney failure in 50s |
| | I.2 | / | / | CKD |
| US2 | III.1 (index) | *UMOD* Sanger sequencing (exons 3,4) | *UMOD* p.T62P (+) | CKD G4 (70-74y), gout (65-69y) |
| | | *MUC1* VNTR sequencing + MS | | |
| | III.2 | *UMOD* Sanger sequencing (exons 3,4) | *UMOD* p.T62P (+) | CKD G5 (55-59y) |

| | | | | |
|---|---|---|---|---|
| | | *MUC1* VNTR sequencing + MS | | |
| | IV.1 | *UMOD* Sanger sequencing (exons 3,4) | *UMOD* p.T62P (-) | unaffected |
| | III.3 | / | / | CKD G5 |
| | III.5 | / | / | CKD G5 |
| | III.6 | / | / | CKD G5 |
| | II.2 | / | / | CKD G5, diabetes mellitus |
| US3 | III.1 (index) | *UMOD* Sanger sequencing (exons 3,4) | *UMOD* p.T62P (+) | CKD G3a (60-64y) |
| | | *MUC1* VNTR sequencing + MS | | |
| | II.1 | *UMOD* Sanger sequencing (exons 3,4) | *UMOD* p.T62P (+) | CKD G5 (75-79y) |
| | | *MUC1* VNTR sequencing + MS | | |
| | II.3 | *UMOD* Sanger sequencing (exons 3,4) | *UMOD* p.T62P (+) | CKD G5 (70-74y) |
| | | *MUC1* VNTR sequencing + MS | | |
| | II.4 | / | / | CKD G4 |
| UK1 | II.2 (index) | Cystic kidney disease panel | *UMOD* p.T62P (+) | CKD G5 (75-79y), minimal proteinuria |
| | | MLPA *HNF1B* | | family history of gout |
| | | *MUC1* VNTR sequencing + MS | | |
| | II.3 | *UMOD* Sanger sequencing | *UMOD* p.T62P (+) | CKD G3b (75-79y) |
| | II.4 | *UMOD* Sanger sequencing | *UMOD* p.T62P (-) | unaffected |
| UK2 | III.2 (index) | *UMOD* Sanger sequencing | *UMOD* p.T62P (+) | CKD G5 (50-54y), minimal proteinuria |
| | | *MUC1* VNTR sequencing + MS | | |
| | | *REN* Sanger sequencing | | |
| | II.2 | *UMOD* Sanger sequencing | *UMOD* p.T62P (+) | CKD G5 (55-59y) |
| | II.4 | *UMOD* Sanger sequencing | *UMOD* p.T62P (+) | CKD G5 (35-39y), gout |
| | III.4 | *UMOD* Sanger sequencing | *UMOD* p.T62P (+) | CKD G5 (35-39y), gout |
| | III.5 | *UMOD* Sanger sequencing | *UMOD* p.T62P (+) | CKD G5 (40-44y), gout (late 20s) |
| | IV.3 | *UMOD* Sanger sequencing | *UMOD* p.T62P (+) | no CKD (15-19y) |

| | | | | |
|---|---|---|---|---|
| | IV.4 | *UMOD* Sanger sequencing | *UMOD* p.T62P (+) | CKD (15-19y) |
| | IV.1 | *UMOD* Sanger sequencing | *UMOD* p.T62P (-) | unaffected |
| | IV.2 | *UMOD* Sanger sequencing | *UMOD* p.T62P (-) | unaffected |
| | III.3 | *UMOD* Sanger sequencing | *UMOD* p.T62P (-) | unaffected |
| | II.3 | *UMOD* Sanger sequencing | *UMOD* p.T62P (-) | unaffected |
| | I.1 | / | / | CKD G5 (55-59y) |
| | I.3 | / | / | CKD |
| UK3 | III.2 (index) | *UMOD* Sanger sequencing | *UMOD* p.T62P (+) | CKD G2 (40-44y) |
| | | cystic kidney disease panel | *GANAB* VUS c.812C>G (p.S271C) | minimal proteinuria |
| | II.1 | *UMOD* Sanger sequencing | *UMOD* p.T62P (+) | CKD G5 (<40y) |
| | | cystic kidney disease panel | *GANAB* VUS c.812C>G (p.S271C) | gout |
| | | *MUC1* VNTR sequencing + MS | | |
| | II.3 | / | / | CKD G5 (40s) |
| | I.3 | / | / | CKD G5 |
| | IV.1 | *UMOD* Sanger sequencing | *UMOD* p.T62P (-) | unaffected |
| | IV.2 | *UMOD* Sanger sequencing | *UMOD* p.T62P (-) | unaffected |
| IRL1 | III.1 (index) | *UMOD* Sanger sequencing | *UMOD* p.T62P (+) | CKD G3b (50-54y), no proteinuria, no hematuria |
| | | *MUC1* VNTR sequencing + MS | | |
| | III.4 | *UMOD* Sanger sequencing | *UMOD* p.T62P (+) | CKD G5 (30-34y) |
| | | *MUC1* VNTR sequencing + MS | | |
| | II.6 | *UMOD* Sanger sequencing | *UMOD* p.T62P (+) | CKD G5 (65-69y), non-enlarged multicystic kidneys |
| | II.7 | *UMOD* Sanger sequencing | *UMOD* p.T62P (+) | CKD G3a (70-74y) |
| IRL2 | III.1 (index) | *UMOD* Sanger sequencing | *UMOD* p.T62P (+) | CKD G5 (70-74y), gout (30-34y), protein 2+, no hematuria |

| | | | | |
|---|---|---|---|---|
| | | *MUC1* VNTR sequencing + MS | | |
| IRL3 | III.1 (index) | *UMOD* Sanger sequencing | *UMOD* p.T62P (+) | CKD G4 (45-49y), no proteinuria, no hematuria |
| | | *MUC1* VNTR sequencing + MS | | |
| | II.3 | / | / | CKD G5 (60-64y), gout, hypertension |
| GE1 | III.1 (index) | *UMOD* Sanger sequencing | *UMOD* p.T62P (+) | CKD G5 (75-79y), no gout, no proteinuria, small kidneys, IF/TA |
| | | *MUC1* SnaPshot Miniseq | | |
| | | *HNF1B* (including MLPA), *REN, SEC61A1* | | |
| | | *COL4A3, COL4A4, COL4A5* | | |
| | III.2 | *UMOD* Sanger sequencing | *UMOD* p.T62P (+) | CKD G5 (70-74y), small kidneys |
| | III.3 | *UMOD* Sanger sequencing | *UMOD* p.T62P (+) | CKD G3a (70-74y) |
| | | *MUC1* SnaPshot Miniseq | | |
| | | *HNF1B* (including MLPA), *REN, SEC61A1* | | |
| | III.4 | *UMOD* Sanger sequencing | *UMOD* p.T62P (-) | unaffected (65-69y) |
| | III.5 | *UMOD* Sanger sequencing | *UMOD* p.T62P (-) | Unaffected (80-84y) |
| | II.1 | / | / | CKD G5 (65-69y) |
| | I.1 | / | / | CKD G5 (65-69y) |
| GEL1 | II.1 (index) | whole genome | *UMOD* p.T62P (+)(tier2), no tier1/2 | CKD G5, Dialysis (Y841, 40-44y), arterial hypertension |
| | II.2 | whole genome | *UMOD* p.T62P (+), no tier1/2 | CKD G5 (N185, 55-59y) |
| | II.3 | whole genome | *UMOD* p.T62P (+), no tier1/2 | CKD non-G5 (65-69y), gout, arterial hypertension |
| GEL2 | II.1 (index) | whole genome | *UMOD* p.T62P (+)(tier2), no tier1/2 | CKD G5, Peritoneal dialysis (X402, 60-64y) |

| | | | | |
|---|---|---|---|---|
| | | | | mild proteinuria, micro. hematuria<br>arterial hypertension |
| GEL3 | II.1 (index) | whole genome | *UMOD* p.T62P (+)(tier2), no tier1/2 | CKD G5 (50-54y), arterial hypertension |
| GEL4 | II.1 (index) | whole genome | *UMOD* p.T62P (+)(tier2), no tier1/2 | multiple small medullary cysts<br>multiple glomerular cysts |
| | I.1 (1969) | whole genome | *UMOD* p.T62P (-) | unaffected |
| GEL5 | II.1 (index) | whole genome | *UMOD* p.T62P (+)(tier2), no tier1/2 | CKD G5 (N185, 65-69y), multiple renal cysts, enlarged kidneys<br>hypertension |

(+) denotes presence and (-) absence of p.Thr62Pro heterozygous change. Tubulopathy gene panel comprising 37 genes, as previously described (9). The cystic kidney disease panel comprises 9 cystogenes (*PKD1, PKD2, GANAB, DNAJB11, HNF1b, PKHD1, UMOD, SEC63, PRKCSH*), as previously described (11). *MUC1* SnaPshot minisequencing as previously described (7). Family GE1 has been previously described as “family 3” (7). *MUC1, HNF1B, REN* and *SEC61A1* can also cause autosomal dominant tubulointerstitial kidney disease, *MUC1* and *UMOD* being by far the most prevalent etiologies (3, 62). Abbreviations as follows: CKD, chronic kidney disease; FEUA, fractional excretion of uric acid; MLPA, multiplex ligation-dependent probe amplification; MS, mass spectrometry; VNTR, variable number tandem repeat; VUS, variant of unknown significance.

**Table S5. Phenotypic categories of all *UMOD* p.Thr62Pro carriers in Genomics England 100,000 Genomes Project.**

| Characteristics of *UMOD* p.Thr62Pro carriers in 100,000 Genomes Project | |
|---|---|
| affected/unaffected | n=69/52 (total=121) |
| phenotypes | Unexplained kidney failure in young people (n=5/N=3) * |
| | Cystic kidney disease (n=5/N=5) ** |
| | CAKUT (n=2/N=2) *** |
| | Brain channelopathy (n=2/N=1) |
| | Charcot-Marie-Tooth disease (n=3/N=2) |
| | Classical tuberous sclerosis (n=2/N=2) |
| | Complex parkinsonism (n=2/N=2) |
| | Congenital myopathy (n=1/N=1) |
| | Dilated Cardiomyopathy (n=1/N=1) |
| | Distal myopathies (n=2/N=2) |
| | Early onset and familial Parkinson's disease (n=1/N=1) |
| | Early onset dementia (n=2/N=2) |
| | Early onset dystonia (n=1/N=1) |
| | Epilepsia plus other features (n=2/N=2) |
| | Epileptic encephalopathy (n=1/N=1) |
| | Familial breast and/or ovarian cancer (n=1/N=1) |
| | Fetal stuctural CNS abnormalities (n=1/N=1) |
| | Hereditary ataxia (n=2/N=2) |
| | Spastic paraplegia (n=2/N=1) |
| | Macular dystrophy (n=3/N=2) |
| | Inherited optic neuropathies (n=1/N=1) |
| | Intellectual disability (n=13/N=13) |
| | Lipoedema disease (n=1/N=1) |
| | Multiple epiphyseal dysplasia (n=1/N=1) |
| | Multiple Tumors (n=1/N=1) |
| | Neurofibromatosis type 1 (n=1/N=1) |
| | Primary immunodeficiency (n=4/N=3) |
| | Rod cone dystrophy (n=1/N=1) |
| | Stickler syndrome (n=1/N=1) |
| | Ultrarare undescribed monogenic disorder (n=4/N=3) |
| | N/A (n=52/N=51) |

* all 3 families appear in Fig S6 (GEL1, GEL2, GEL3)
** 2 families appear in Fig S6 (GEL4, GEL5), other 3 families have mutations in *PKD1/2*
*** Phenotype not consistent with UMOD pathophysiology

**Table S6. Characteristics of CKD case and control groups in the 100,000 Genomes project.**

| | No CKD | CKD any stage | No renal failure | Renal failure | All T62P heterozygotes | T62P heterozygotes with CKD | T62P heterozygotes without CKD |
|---|---|---|---|---|---|---|---|
| n (total) | 84,465 | 4,453 | 87,567 | 1,351 | 120 | 11 | 109 |
| n probands | 31,094 (36.8%) | 2,235 (50.2%) | 32,371 (37.0%) | 958 (70.9%) | 57 (47.5%) | 6 (54.6%) | 51 (46.8%) |
| Age (years, mean±SD) | 43.6±21.6 | 56.7±24.9 | 44.1±22.0 | 49.0±21.0 | 43.0±19.7 | 57.5±17.8 | 41.5±19.4 |
| Females | 53.6% | 47.0% | 53.4% | 43.2% | 58.3% | 18.2% | 62.4% |
| Ethnicity: Asian or Asian British | 8.1% | 6.6% | 8.0% | 9.0% | 0.8% | 9.1% | 0% |
| Ethnicity: Black or Black British | 2.2% | 4.1% | 2.2% | 7.1% | 0% | 0% | 0% |
| Ethnicity: Mixed | 1.8% | 1.6% | 1.8% | 1.6% | 0.8% | 0% | 0.9% |
| Other ethnic groups | 1.7% | 1.4% | 1.7% | 1.8% | 0.8% | 0% | 0.9% |
| Ethnicity: White | 68.8% | 68.6% | 68.8% | 66.3% | 80.1% | 54.5% | 82.6% |
| Ethnicity not stated or not available | 17.4% | 17.7% | 17.5% | 14.2% | 17.5% | 36.4% | 15.6% |
| Probands with affected parent(s) | 15.1% | 20.8% | 15.2% | 24.5% | 22.8% | 50.0% | 9.2% |
| Probands with affected sib(s) | 14.5% | 18.6% | 14.6% | 20.8% | 15.8% | 33.3% | 6.4% |

Information on age, gender and ethnicity provided for all individuals recruited (probands + relatives). Gender information and ethnic categories as provided by the 100,000 Genomes Project. CKD, chronic kidney disease.

**Table S7. Urinary UMOD levels in controls, *UMOD* p.Thr62Pro carriers and ADTKD patients with canonical *UMOD* mutations.**

| Fam./cohort | Patient/n | eGFR (mL/min/1.73m$^2$) mean±SD | uUMOD (mg/g creat) mean±SD |
|---|---|---|---|
| CH1 | III.1 | 43 | 3.0 |
| CH1 | III.2 | 90 | 8.0 |
| UK3 | III.2 | 60-89 | 13 |
| IRL1 | III.1 | 30-44 | 16 |
| IRL1 | II.6 | <15 | 7 |
| IRL1 | II.7 | 45-59 | 15 |
| GCKD | T62P-1 | 30-45 | 4 |
| GCKD | T62P-2 | 30-45 | 7 |
| GCKD | T62P-3 | 30-45 | 8 |
| GCKD | T62P-4 | 45-60 | 16 |
| GCKD | T62P-5 | >90 | 4 |
| Controls CoLaus (eGFR 30-60) | n=226 | 54.4±5.6 | 17±10 |
| Canonical *UMOD* mutation (eGFR 30-60) | n=26 | 40.6±8.4 | 2±2 |

Abbreviations: CoLaus, Cohorte Lausannoise; GCKD, German Chronic Kidney Disease Cohort; uUMOD, urinary uromodulin.

**Table S8. Overview of participating cohorts/centres and ethics committees/institutional review boards.**

| Cohort name | Ethics Committee / Institutional Review Board (IRB) | approved / waived |
|---|---|---|
| UK Biobank | North West Multi-centre Research Ethics Committee which covers the UK (approval number: 11/NW/0382) | Approved |
| 100,000 Genomes Project | East of England - Cambridge South Research Ethics Committee, UK (REC Ref 14/EE/1112). | Approved |
| German Chronic Kidney Disease Cohort | Deutsches Register Klinischer Studien DRKS00003971, Ethik-Kommission der Friedrich-Alexander-Universität Erlangen-Nürnberg, Germany, Nr 3831 | Approved |
| ADTKD cohort (Belgium/Switzerland/USA) | Institutional review board of Wake Forest School of Medicine, North Carolina, USA (Wake Forest University Health Sciences IRB00000352 “Characteristics of Individuals with Inherited Kidney Disease”) | Approved |
| | Institutional review board of the Université Catholique de Louvain (UCL) Medical School and Saint Luc University Hospital, Belgium (2011/04MAI/184) | Approved |
| | European Community's Seventh Framework Programme “European Consortium for High-Throughput Research in Rare Kidney Diseases (EURenOmics) Ethics Advisory Board. Participating centers (amongst others): University of Zurich (Switzerland), Newcastle University (UK) | Approved |
| Local IRB: UK | The North East - Newcastle & North Tyneside 1 Research Ethics Committee, UK (18/NE/350) | Approved |
| Local IRB: Ireland | Ethics Review Board of Beaumont Hospital, Dublin, Ireland (REC 19/28) | Approved |
| Local IRB: Germany | Ethics committee of the Friedrich-Alexander University Erlangen-Nürnberg, Germany (approval number: 251_18 B) | Approved |
